# Recovery of Dexterous Motor Control via Non-Monosynaptic Corticospinal Pathways

**DOI:** 10.64898/2026.03.24.26348827

**Authors:** Erynn Sorensen, Luigi Borda, Julia Ostrowski, Roberto M. de Freitas, Nikhil Verma, Lee E. Fisher, George F. Wittenberg, Peter Gerszten, Doug J. Weber, Elvira Pirondini, Monica Gorassini, John W. Krakauer, Marco Capogrosso

## Abstract

Fine motor control of the human arm is assumed to depend on monosynaptic connections between the motor cortex and spinal motoneurons. We report that people with post-stroke hemiparesis could regain dexterous control using non-monosynaptic corticospinal tract (CST) projections during epidural cervical spinal cord stimulation (SCS). Participants in our pilot clinical study demonstrated the ability to improve strength, reaching smoothness, and fine force control of the arm and hand while receiving continuous stimulation. Detailed electrophysiology of corticospinal connectivity with transcranial magnetic stimulation, electromyography spectral analysis, and single motor-unit firing showed that SCS did not reliably strengthen CST-activation of motoneurons. Instead, residual CST axons sculpted the strength of spinal reflexes produced by each pulse of SCS via polysynaptic mechanisms, including presynaptic gating of sensory afferent excitability, to promote functionally relevant muscle activation patterns. Our findings reveal the ability of non-monosynaptic CST pathways to finely tune spinal motor output in humans after stroke.

## INTRODUCTION

It is well established that evolutionary divergence led to the development of the corticospinal tract (CST): a neural pathway that supports skilled use of the arm and hand^1–6^. Humans, in particular, have evolved monosynaptic connections of the CST to spinal motoneurons that are largely absent in other mammals, providing a direct substrate for fine motor control^1–6^. Today, it is often assumed that this monosynaptic component is necessary for dexterous control of the arm and hand^1,4–6^. Indeed, humans are extremely vulnerable to CST damage, and even small lesions in the internal capsule lead to permanent loss of finger, hand, and overall upper limb dexterity^7–10^, here defined to encompass not only the ability to individuate fingers, but also the ability to accurately target and grade forces of the arm and hand and to perform smooth movements^11,12^. For this reason, the presence of motor evoked potentials (MEPs) elicited by transcranial magnetic stimulation (TMS) of the motor cortex, a biomarker of CST integrity and associated to monosynaptic connections, is assumed as a predictor for stroke recovery^10,13–15^.

However, mounting evidence suggests that non-monosynaptic collaterals of CST axons play a significant role in skilled function^2,3,5,16–20^ raising questions on the relevance of these pathways in humans and more specifically on whether monosynaptic CST connections are truly necessary to recover dexterous movements after a lesion.

Assuming this was the case, we explored whether neurostimulation could support descending monosynaptic neural signals from a damaged CST in recruiting spinal motoneurons, thereby enabling recovery of upper-limb dexterity. We specifically focused on epidural cervical spinal cord stimulation (SCS) which facilitates volitional control of the arm and hand in humans with spinal and subcortical brain lesions^21–32^. By stimulating glutamatergic sensory afferents in the dorsal roots, SCS excites spinal motoneurons, improving the ability of residual CST axons to depolarize motoneurons and produce functional muscle contractions after a lesion^26,27,33–41^. Building on this evidence, we tested the hypothesis that SCS could improve CST function, and therefore upper-limb dexterity, in people with CST damage.

To do so, we conducted a series of experiments within an exploratory clinical trial (NCT04512690) of SCS in humans with chronic post-stroke hemiparesis^23,24^ and we found that participants could immediately improve precise force control of the arm and hand, as well as reaching smoothness and muscle coordination, when SCS was turned on. Based on current models of human motor control, we assumed SCS was facilitating monosynaptic CST-activation of spinal motoneurons to enable these dexterous movements. To test for this, we performed a series of behavioral and electrophysiological experiments. Surprisingly, we did not find conclusive evidence that SCS functionally facilitates activation of motoneurons via the monosynaptic CST. Instead, our cumulative results suggested that residual CST projections regulated the excitatory drive of SCS to spinal motoneurons via presynaptic gating of primary afferent excitability^42–44^. Volitional regulation through these non-monosynaptic CST pathways effectively steered the excitatory drive of SCS towards functionally relevant muscles, thereby re-establishing partial supraspinal control of arm and hand motoneurons. Through this mechanism, CST inputs without strong monosynaptic connections to motoneurons contributed significantly to dexterous motor recovery, revealing their previously underappreciated capacity to support skilled arm and hand control.

## RESULTS

### SCS improves fine motor control in humans post-stroke

Our experiments were executed in the context of a clinical trial (NCT04512690) testing preliminary safety and efficacy of SCS to improve function in people with post-stroke hemiparesis^23,24^. Briefly, participants were implanted with epidural leads stimulating the dorsal roots innervating muscles of the arm and hand (C4–T1). We identified patient-specific stimulation patterns that promoted muscle activation (Figure 1A, see methods) and measured changes in motor control that occurred when SCS was turned on^23,24^.

**Figure 1.**
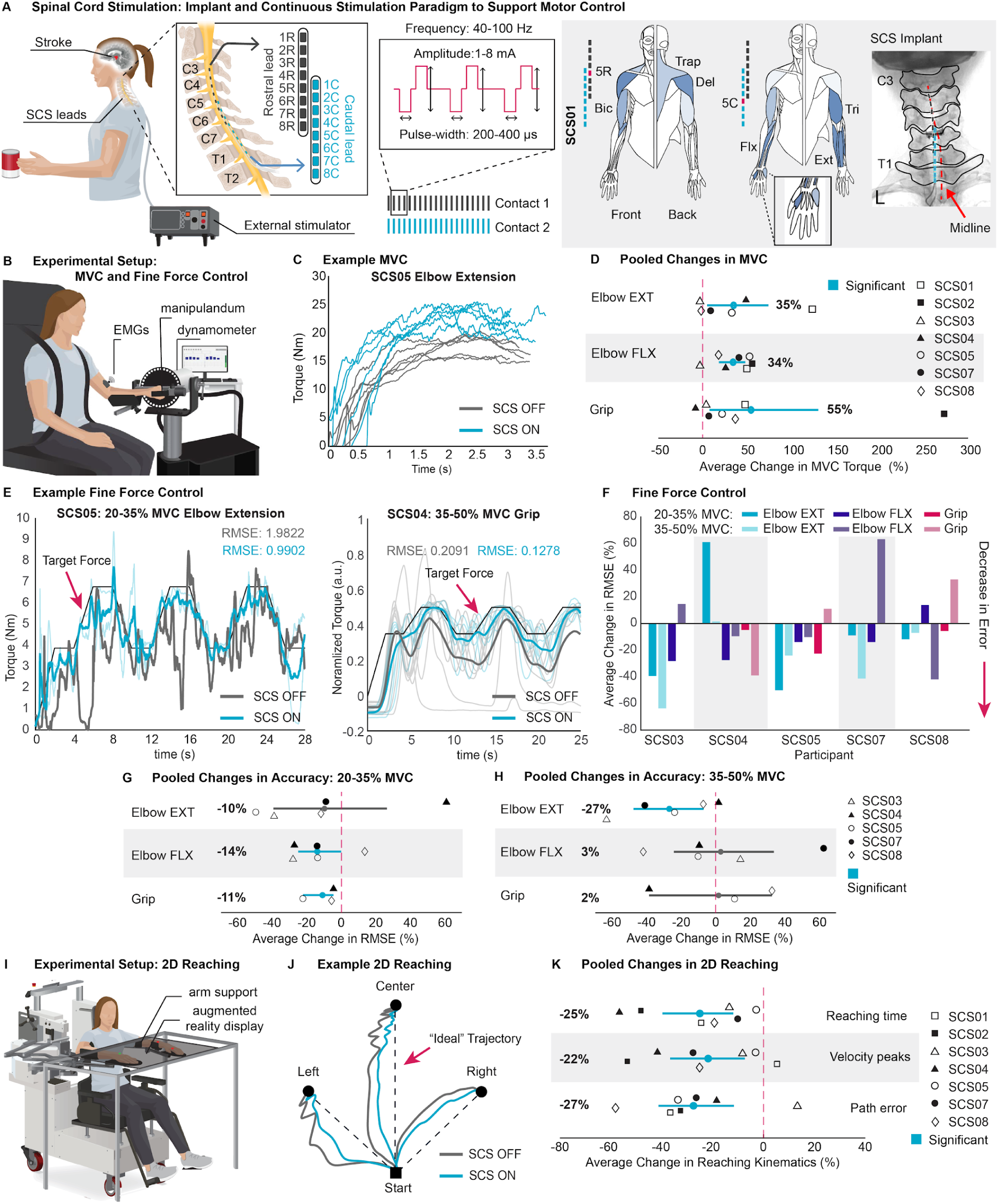
SCS improves fine motor control in humans post-stroke. **(A)** Illustration of SCS applied via two 8-contact epidural leads connected to an external multi-channel stimulator. Shown in the grey box is a representative recruitment map of normalized muscle activation from intraoperative recordings in SCS01 (white=0; dark blue=1) for selected contacts (pink labeled). X-ray shows rostral (blue) and caudal (grey) lead positions relative to midline (pink). **(B)** Experimental setup for panels C–H. (*Not shown*: handheld dynamometer used to record grip force). **(C)** Example torque traces during MVC elbow extension (SCS05). SCS OFF (grey), SCS ON (blue), n=6 per condition. **(D)** Group-level changes in MVC torque for elbow extension, flexion, and grip. Percent differences calculated per participant between mean SCS ON and SCS OFF (n=5–6 per condition) and bootstrapped to obtain 95% confidence intervals (CI). **(E)** Example mean force traces versus target during elbow extension (SCS05) and grip (SCS04). Individual repetitions shown with reduced opacity. Left: 20–35% MVC extension (SCS OFF (grey) n=1, SCS ON (blue) n=2). Right: 35–50% grip (SCS OFF (grey) n=9, SCS ON (blue) n=6). Mean root-mean-square error (RMSE) from target written in corresponding colors. **(F)** Mean percent differences in RMSE during fine force control between SCS ON and SCS OFF per participant. Darker shades: 20–35% MVC; lighter shades: 35–50% MVC. Blue: elbow extension; purple: elbow flexion; pink: grip. Negative values indicate decreased error with SCS ON. **(G–H)** Group-level changes in force control accuracy during (G) 20–35% and (H) 35–50% MVC. Percent differences were calculated per participant between mean SCS ON and SCS OFF (extension: n=1–3, flexion: n=1–3, grip: n=6–9 per condition) and bootstrapped to obtain 95% CIs. **(I)** Experimental setup for panels J–K. **(J)** SCS04 average hand trajectory during 2D reaching. SCS OFF (grey), SCS ON (blue). **(K)** Group-level changes in reaching time, velocity peaks, and path error. Percent differences were calculated between mean SCS ON and SCS OFF (n=14–30 per condition) and bootstrapped to calculate 95% CIs. *Panels D, G, H, and K: Participant-level mean percent differences were resampled with replacement (10,000 iterations) to estimate the group-level mean and 95% CI. CIs excluding zero (blue) indicate significant changes; the mean percent change is shown for each interval*.

Assuming that SCS facilitates residual descending inputs^26,27,37,39^, we expected increases in strength with SCS. We compared torque during isometric maximum voluntary contractions (MVC) with SCS on versus off (Figure 1B). When SCS was on, participants immediately increased maximum torque production by an average of 35% during elbow extension, 34% during elbow flexion, and 55% during grip (Figure 1D, see example 1C).

While necessary, strength is not the only component of motor control. To independently assess dexterous improvements from changes in gross strength, we developed a force modulation protocol using the same isometric setup as MVC, but instead designed to measure graded force control^7,11,12^. In this task, participants were asked to finely modulate their force output between two difficulty ranges, 20–35% (low) and 35–50% (mid) of their MVC capacity, as assessed with SCS off. Figure 1E exemplifies this task. Improvements in force-tracing error were common across participants, with the majority of tested tasks demonstrating decreases in error (Figure 1F), albeit with differences per joint and force level. Across-patient significant improvements occurred during mid-range elbow extension (Mean: −27%), low-range elbow flexion (Mean: −14%), and during low-range grip (Mean: −11%) (Figures 1G,1H). Interestingly, increases in accuracy were not solely explained by increases in agonist muscle activity as several participants demonstrated an overall decrease in muscle activity (S1A–B). Instead, the fine force control task revealed a trend towards rebalancing agonist/antagonist muscle ratios (S1C–D).

We additionally implemented a two-dimensional center-out reaching task wherein participants were asked to move, as naturally as possible, from a central target towards peripheral targets on the same plane (Figure 1I). Performance was quantified by time-to-target, movement smoothness (velocity peaks), and path efficiency (deviation from an ideal straight trajectory) in both SCS off and on conditions. An example of this task can be seen in Figure 1J. We saw significant decreases in reaching time (Mean: −25%), velocity peaks (Mean: −22%), and pathing error (Mean: −27%) across all participants (Figure 1K). This was supported by a clear increase in triceps EMG activity (Mean: 52%) along with a contrasting decrease in brachioradialis activity (Mean: −21%) with SCS on. There was a strong decoupling of the triceps and brachioradialis during reaching (Mean: 91%) (S1F–G). Together, these results suggest that SCS not only enhances strength, but also improves participants’ ability to grade and coordinate forces of the arm and hand to produce smoother, more accurate, and faster dexterous movements.

### SCS improves cortico-muscular coupling

If SCS is facilitating CST inputs, then this should be reflected by a change in intermuscular coherence within the EMG beta-band, which is a measure of the strength of motor-cortical common synaptic input to two muscles^45–51^. We quantified beta-band intermuscular coherence between two agonist muscles (biceps and brachioradialis) and two antagonist muscles (biceps and triceps) during force modulation (Figure 2A), specifically during low-range elbow flexion. Agonist muscle pairs demonstrated increased coherence within the beta-band with SCS on (Figure 2B,2D). Contrastingly, antagonist muscle pairs showed either no change or a decrease in coherence (Figure 2C,2D). Thus, SCS selectively facilitates cortico-muscular coupling of muscles agonistic to a task, while having no effect or even decreasing cortico-muscular coupling of unwanted antagonistic muscles. This suggests that SCS specifically facilitates supraspinal recruitment or coordination of muscles relevant for dexterous control, likely via the CST, which is compatible with improved monosynaptic function^49^.

**Figure 2.**
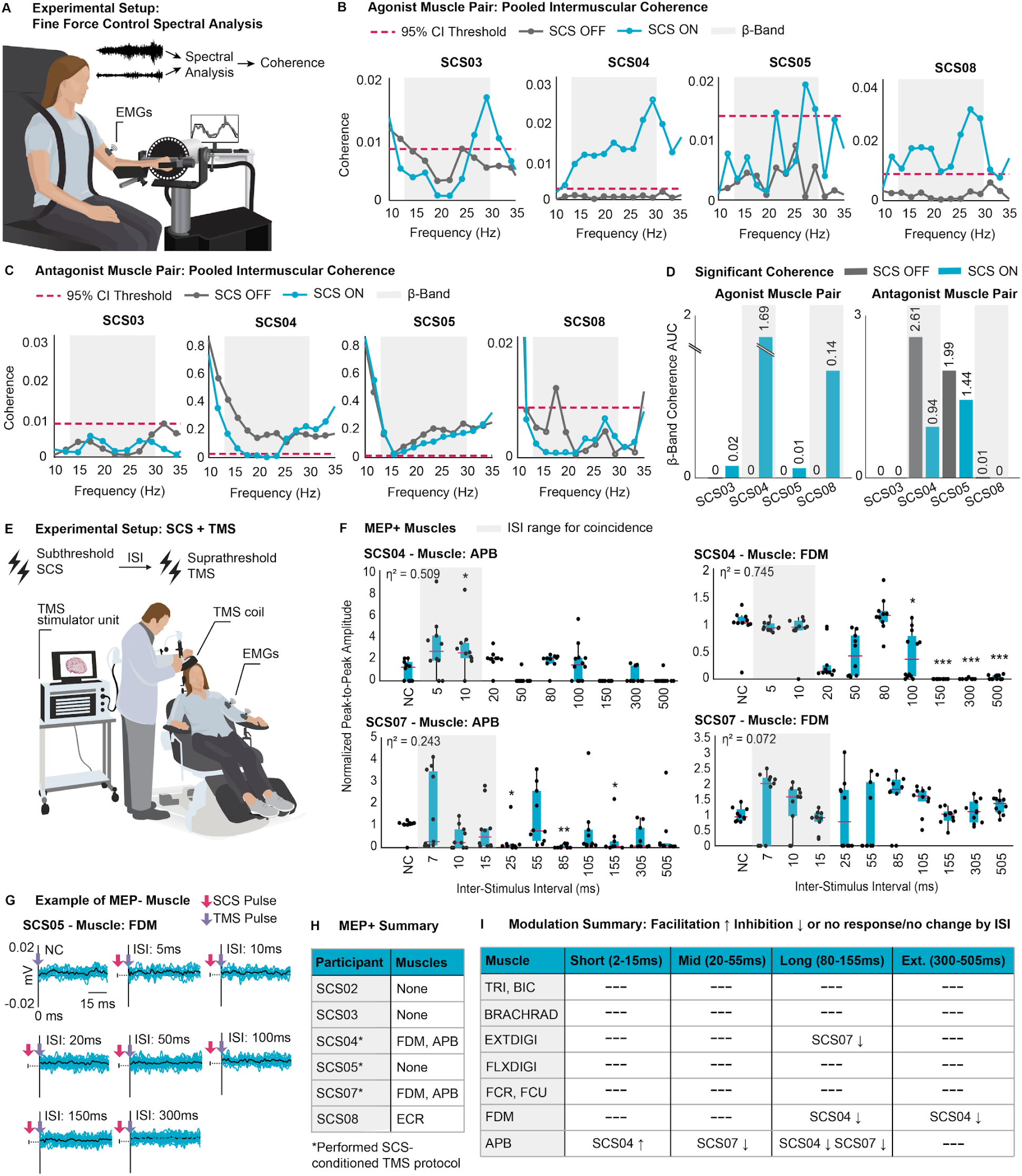
SCS improves cortico-muscular coupling, but not by facilitating corticospinal-motoneuron connections. **(A)** Experimental setup for panels B–D; 20–35% MVC elbow flexion fine-force control task. **(B–C) (B)** Agonist (Biceps-Brachioradialis) and **(C)** Antagonist (Biceps-Triceps) muscle pairs: pooled intermuscular coherence; SCS OFF (grey), SCS ON (blue) (n=1–3 per condition). Shaded region indicates beta-band (13–30hz). Dashed red line represents 95% confidence threshold for true coherence, computed using the pooled number of independent spectral segments across trials. **(D)** Beta-band area-under-the-curve (AUC) for pooled coherence above the 95% confidence threshold. Bars show AUC for SCS OFF (grey) and SCS ON (blue). Higher values in agonist and lower values in antagonist muscle pairs under SCS ON indicate stronger functional cortico-muscular coupling. **(E)** Experimental setup for panels F–I. Single pulses of subthreshold SCS delivered prior to single pulses of suprathreshold TMS at varying inter-stimulus-intervals (ISI). **(F)** MEP-positive muscles across the participants who underwent this protocol (SCS04, SCS05, SCS07). Boxplots (median ± IQR, whiskers = 1.5×IQR) show peak-to-peak conditioned MEP amplitudes normalized to the no-conditioning (NC) MEP across ISIs (2–505ms) with overlaid dots (n=10 per ISI). Grayed regions indicate where EPSPs from SCS and TMS would be expected to coincide at the motoneuron. *Kruskal–Wallis with Dunnett-type post hoc versus NC; η*^*2*^ *effect size. *p<0*.*05, **p<0*.*01, ***p<0*.*001*. **(G)** Representative TMS-triggered responses at varying ISIs from an MEP-negative muscle (SCS05: FDM). No responses and no facilitation shown. Individual responses (blue); mean response (black). **(H)** Summary of muscle MEP status by participant. **(I)** Summary of MEP modulation across the 3 participants who underwent this protocol (SCS04, SCS05, SCS07). Arrows indicate facilitation (↑) or inhibition (↓); dashes = no significant modulation or no response. *TRI: tricep, BIC: bicep, BRACHRAD: brachioradialis, EXTDIGI: extensor digitorum, FLXDIGI: flexor digitorum, ECR: extensor carpi radialis, FCR: flexor carpi radialis, FCU: flexor carpi ulnaris, FDM: first dorsal interosseous, APB: abductor pollicis brevis*.

### SCS does not reliably facilitate CST connections to motoneurons

SCS is known to directly recruit sensory afferents that provide excitatory input to spinal motoneurons^33–35,38,41^. This is thought to bring motoneuronal membrane potentials closer to firing threshold and make them more responsive to supraspinal input^26,27,33,37,39^, which could explain our observed increase in cortico-muscular coupling. Similarly, we expect a single excitatory SCS pulse to lower the effective motoneuron threshold for the duration of the elicited compound excitatory postsynaptic potential (EPSP), enabling the monosynaptic CST to depolarize motoneurons more readily. To test this we delivered a subthreshold SCS pulse to condition motoneuronal membranes, followed by a single pulse of TMS over the motor cortex (Figure 2E) which is known to activate CST projections to elicit MEPs^52–54^. We expected that by improving monosynaptic CST function, conditioning with SCS would lead to significant facilitation of the MEPs. Specifically, if an MEP was already present at rest, conditioning should increase its peak-to-peak amplitude; if no MEP was present, conditioning could enable its emergence. So long as TMS is delivered within approximately 20ms post-SCS trigger, we would expect to see coincidental arrival of EPSPs from the cortex (TMS) and the dorsal roots (SCS)^14,55^. This was not what we observed.

Across the three tested participants, if a muscle was MEP-negative, SCS never led to the generation of an MEP (Figure 2G). It is worth noting that the majority of muscles across participants were MEP-negative (Figure 2H). For the four muscles across participants which were MEP-positive, there was no consistent facilitation of MEPs. Indeed, only SCS04 showed facilitation of the abductor pollicis brevis when the inter-stimulus-interval (ISI) was 10ms (Figure 2F, 2I). Although, subthreshold monosynaptic activity may still have been present, the overwhelming lack of MEP facilitation casts doubts on our leading hypothesis and suggests that improvements in dexterity and corticomuscular coherence may be explained by other mechanisms rather than improvement of monosynaptic CST function during SCS.

### Residual CST connections presynaptically facilitate spinal reflex pathways

If SCS does not directly facilitate CST connections to motoneurons, how else might we explain increased cortico-muscular coupling with SCS? Our recent work has shown that residual supraspinal inputs transform otherwise subthreshold SCS-induced EPSPs into action potentials by modulating the membrane potential of motoneurons. In this view, SCS entrains motoneuron firing but does not directly trigger action potentials; instead residual supraspinal input determines which SCS pulses are transformed into action potentials^33^. This reframes the interaction: SCS remains the primary driver of motoneuron recruitment, while supraspinal input modulates when it becomes effective. As a corollary to this, sub-threshold descending input could sculpt the strength of motoneuron activation by SCS to facilitate activation of upper limb muscles. This framework provides a mechanistic basis for our next experiment, in which we did the reverse protocol and used a single pulse of subthreshold TMS to bias motoneurons and then delivered a single pulse of suprathreshold SCS to test if residual corticospinal inputs can facilitate SCS-evoked responses (Figure 3A).

**Figure 3.**
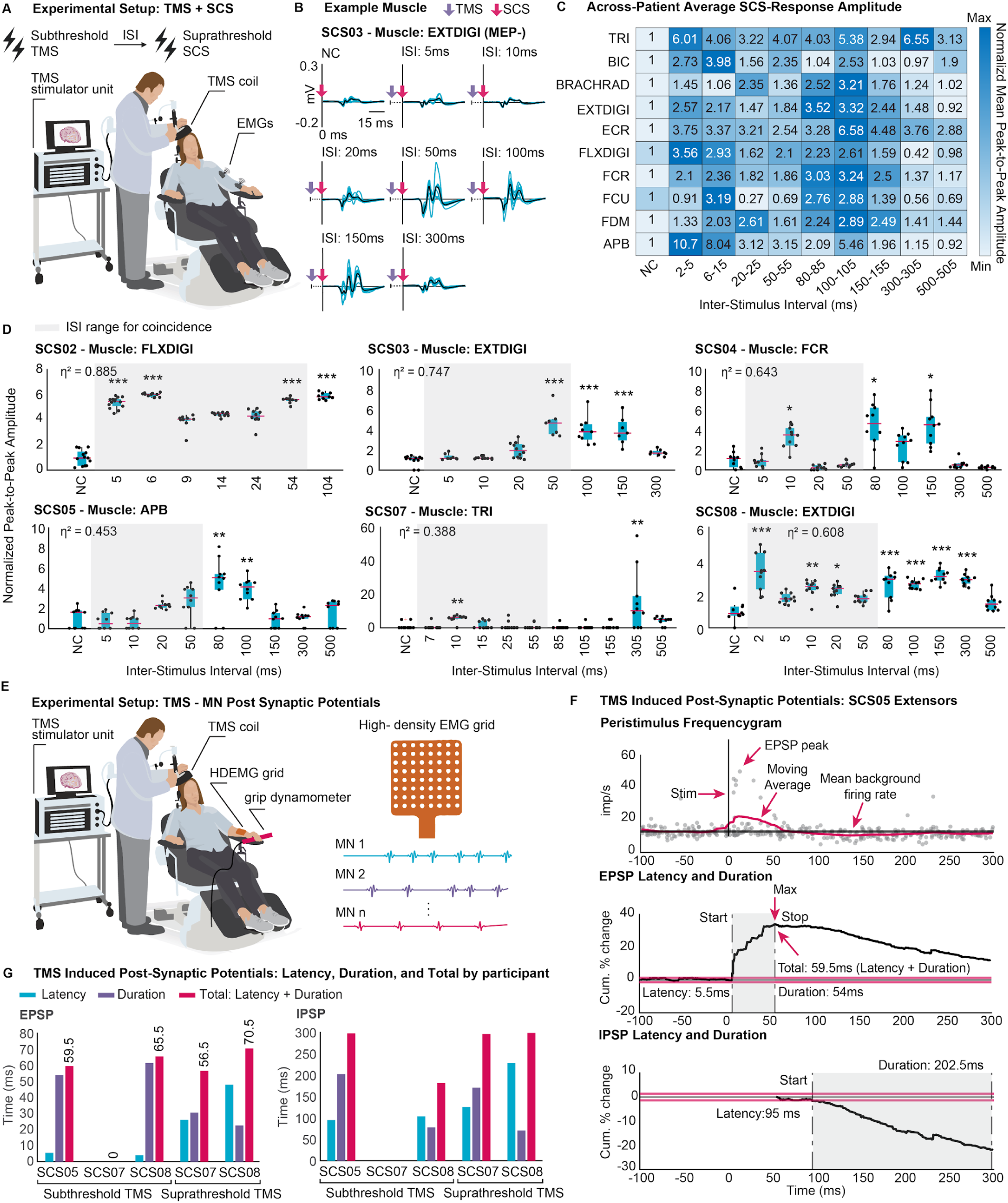
Residual CST connections presynaptically facilitate spinal reflex pathways. **(A)** Experimental setup for panels B–D. Single pulses of subthreshold TMS delivered prior to single pulses of suprathreshold SCS at varying inter-stimulus-intervals (ISI). **(B)** Representative SCS-triggered responses at varying ISIs from an MEP-negative muscle (SCS03: EXT DIGI). Strong facilitation from no-conditioning (NC) baseline at ISIs 50–150ms. Individual responses (blue); mean response (black). **(C)** Across-participant average response amplitude normalized to each muscle’s response in the no-conditioning (NC) baseline. Heatmap shows mean normalized peak-to-peak amplitudes per binned ISI and muscle; darker blue indicates larger amplitude. Two groupings appear: a short ISI facilitation (2–15ms) and a long ISI facilitation (80–155ms). **(D)** Example muscle from each participant under the TMS+SCS conditioning protocol. All participants demonstrate a long ISI facilitation. Boxplots (median ± IQR, whiskers = 1.5×IQR) show normalized peak-to-peak SCS-evoked response amplitudes across ISIs (2–505ms) with overlaid dots (n=8–14 per ISI). Grayed regions indicate where EPSPs from SCS and TMS would be expected to coincide at the motoneuron. *Kruskal–Wallis with Dunnett-type post hoc versus NC; η*^*2*^ *effect size. *p<0*.*05, **p<0*.*01, ***p<0*.*001*. **(E)** Experimental setup for assessing TMS-induced postsynaptic potentials in motoneurons. Sub- and supra-threshold TMS delivered (n≈80 per condition) every 5 seconds while HDEMG recorded motor-unit firing in the forearm during a low-level contraction (20% MVC grip). **(F)** Example participant (SCS05, subthreshold TMS, extensors, single motor unit). Top: peristimulus frequencygram (PSF) showing changes in motor-unit firing probability aligned to TMS onset (grey dots: individual instantaneous firing rates; pink: moving average; black: mean background firing rate). Middle and bottom: cumulative sum analysis illustrating excitatory (EPSP) and inhibitory (IPSP) postsynaptic potentials measuring latency (passing of baseline threshold firing) and duration (maximum or minimum in the cumulative sum trace). Pink bounding boxes: baseline thresholds; black: cumulative sum trace; shaded region: EPSP or IPSP period. **(G)** Summary of EPSP and IPSP characterizations across participants. Bar plots show latency (blue), duration (purple), and total time till postsynaptic potential end (pink) by patient and condition (n=1–4 motor-units). *TRI: tricep, BIC: bicep, BRACHRAD: brachioradialis, EXTDIGI: extensor digitorum, FLXDIGI: flexor digitorum, ECR: extensor carpi radialis, FCR: flexor carpi radialis, FCU: flexor carpi ulnaris, FDM: first dorsal interosseous, APB: abductor pollicis brevis*.

We found consistent facilitation compared to the no-conditioning (NC) response amplitude at short intervals (~2–15ms), corresponding to the times at which sensory afferent and CST EPSPs could coincide and summate at the motoneurons. Surprisingly, we also observed strong facilitation at long intervals (~80–155ms) (Figure 3C-D), far outside the expected window for summation. This long interval facilitation was often substantially stronger than that at short intervals. This was true regardless of MEP status. An example of this is illustrated in Figure 3B. Indeed, if we sum the normalized mean peak-to-peak reflex amplitudes at each ISI, TMS delivered 100-105ms before SCS led to the strongest facilitation. Furthermore, when the abductor pollicis brevis (a finger muscle with strong monosynaptic corticospinal input that disproportionately contributes to the second-strongest ISI (2–5ms)) is excluded from the summation, facilitation at the 100-105ms ISI becomes even more dominant (Figure 3C). We controlled for excitation arising from arousal-related reticulospinal inputs evoked by the sound of the TMS “click”^56^. Across the sham-tested participants, only 12.5% of recorded muscles (3 muscles) responded to this control, as opposed to the 54% of muscles which were facilitated by TMS (S2B). This shows that while reticulospinal-arousal may have played some role, it does not explain our TMS-conditioning results.

Interestingly, the timings of long-interval facilitation matched with previously studied facilitation of H-reflexes by TMS pulses, which have been proposed to reflect CST-induced primary afferent depolarization (PAD)^42–44^. This suggests that the CST could facilitate the transmission of SCS to motoneurons via presynaptic modulation of Ia-afferent excitability. However, in stroke, corticospinal EPSPs may be temporally dispersed or prolonged, causing them to arrive at significantly longer latencies than expected^53,57,58^ which could be an alternative explanation for the observed long-interval facilitation.

To rule out the argument of unexpectedly long EPSPs, we recorded single motor-unit activity during an isometric hold. Changes in instantaneous firing rate upon the application of TMS are known to be directly proportional to membrane potential fluctuations, allowing visualization of EPSPs noninvasively with a peristimulus frequencygram (PSF)^59–61^. We used the cumulative sum of this increase in instantaneous firing to estimate the time at which EPSPs ended. For example, in SCS05 the cumulative sum of the PSF peaks at t=59.5ms indicating that the instantaneous firing rate returned to baseline. Combining both visual information and quantitative analysis of the cumulative firing across participants, we can estimate that TMS-induced EPSPs ended approximately 63ms post-TMS trigger (Figure 3G,S2C–D).

The finding that EPSPs ended on average 63ms post-TMS trigger (well before the observed long-interval facilitation 80–155ms) and that motoneurons firing rates remained flat, if not inhibited afterwards, indicate that the observed facilitation of SCS-induced reflexes at long ISIs must be occurring pre-synaptically to the motoneurons. This aligns with the hypothesis that SCS reflexes are facilitated via CST-induced PAD, which has a long time-course of 100–200ms^42^.

### Volitional modulation of post-activation depression demonstrates presynaptic control of sensory inputs

We aimed to further confirm that the CST presynaptically modulates the transmission of SCS, and to establish PAD as a potential presynaptic mechanism by which volition (not induced by TMS) can facilitate reflex strength.

For this we studied the effects of volitional muscle activation on post-activation depression: i.e. a reduction in motoneuronal responses following repeated activation of the sensory afferents. Briefly, when motoneurons are activated via reflex pathways (as with SCS) the amplitude of evoked muscle potentials decreases over time with rates proportional to the frequency of stimulation. This is thought to be a presynaptic effect related to the amount of neurotransmitter release at the synapses which cannot replenish vesicles fast enough to follow each action potential. This causes a progressive (exponential) drop in neurotransmitter release for each consecutive pulse after the first^38,62–64^. By measuring the change in rate of exponential decay during a train of SCS we could directly assess the ability of volition to presynaptically modulate the transmission of SCS (Figure 4A–C).

**Figure 4.**
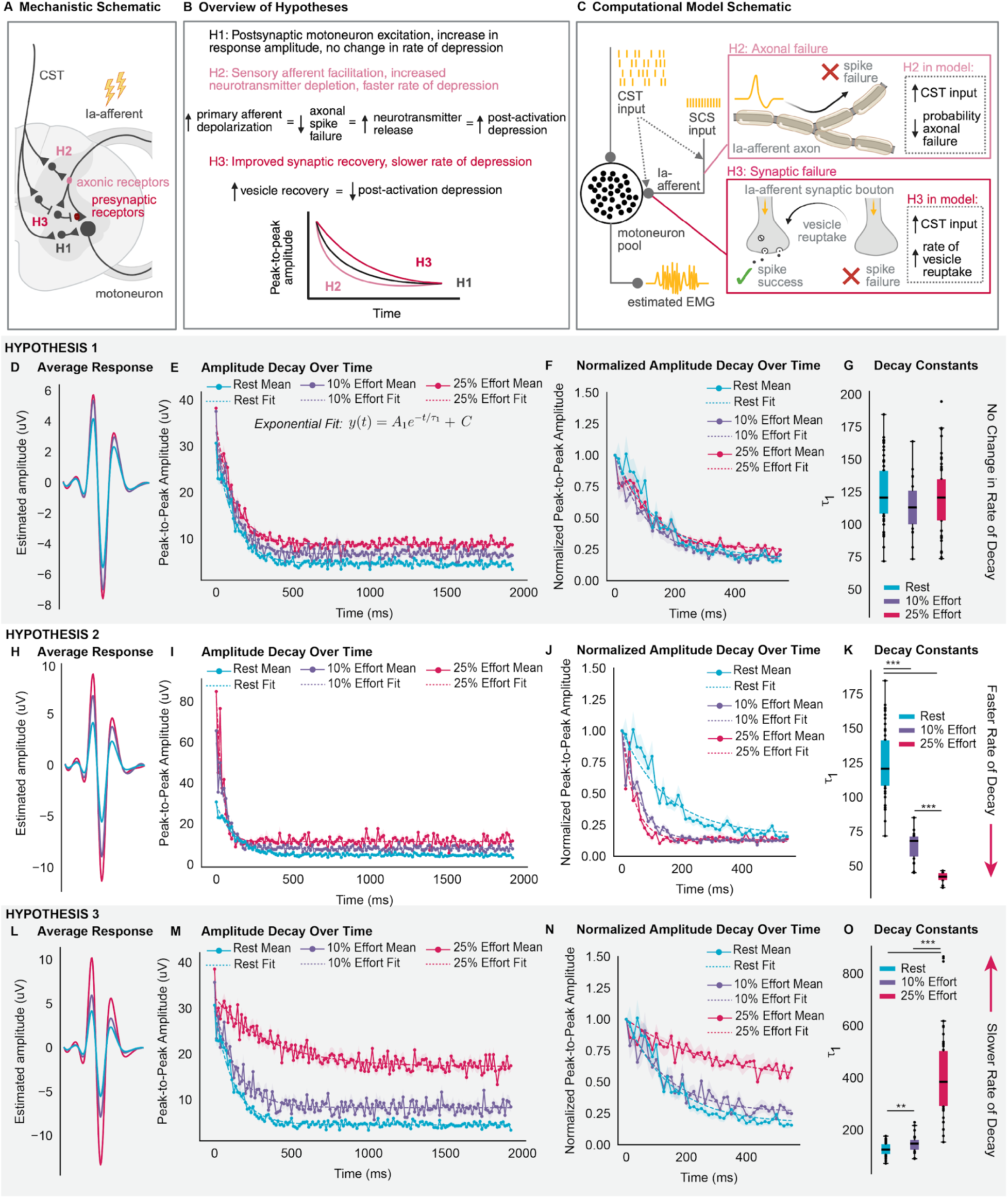
In silico: volitional modulation of post-activation depression demonstrates presynaptic control of sensory inputs. **(A–C)** Mechanistic and computational models illustrating how CST inputs may modulate sensory afferent transmission. The model simulates la-afferent activation by SCS converging with indirect and direct CST input to the motoneuron pool and tests three hypotheses for how the CST could influence SCS transmission. Insets show modeled axonal failure and impaired synaptic recovery mechanisms linked to post-activation depression and the predicted decay trajectories for each hypothesis. **(A)** Triangular terminals indicate excitatory neurons. Flat terminals indicate inhibitory neurons. **(D–G)** H1: Model output showing muscular responses during a SCS burst at rest (blue), with 10% (purple), and 25% effort (pink). **(D)** Average waveform across effort levels. **(E)** Peak-to-peak amplitude across burst (mean (solid line) ± SEM; exponential fit (dashed line); n=10–15 bursts). **(F)** Normalized peak-to-peak amplitude across burst (mean (solid line) ± SEM; exponential fit (dashed line); n=10–15 bursts). **(G)** Boxplots (median, IQR, whiskers = 1.5×IQR) summarize fitted τ_1_ values with overlaid dots (n=10–15 bursts). *Kruskal-Wallis with Tukey-Kramer post hoc, *p<0*.*05, **p<0*.*01, ***p<0*.*001. Effect size reported as η*^*2*^. Increased postsynaptic activation of motoneurons with increasing effort yield no change in τ_1_ values, despite larger response amplitudes. **(H–K)** H2: Same format as (D–G). Reduced axonal failure with increasing effort yield smaller τ_1_ values, consistent with greater neurotransmitter depletion. **(L–O)** H3: Same format as (D–G). Shorter vesicle-reuptake time constants with increasing effort yield larger τ_1_ values, consistent with improved synaptic recovery.

For example, if the CST was only exciting motoneurons post-synaptically (H1 Figure 4A), then the rate at which repeated SCS-evoked responses decayed would not change when comparing rest versus volition because the CST would not affect the rate of neurotransmitter release from the afferents. Instead, one would only observe higher peak-to-peak amplitudes due to higher motoneuron excitability (H1). In contrast, if the CST acted presynaptically to induce PAD^42–44^ (H2 Figure 4A), the facilitation of Ia-afferent excitability would result in more neurotransmitter release. This would lead to higher peak-to-peak response amplitudes due to increased transmission. However, this would also accelerate vesicle depletion in the synapses; exacerbating post-activation depression and causing a faster attenuation of SCS-evoked motoneuronal responses (H2). Alternatively, the CST could be acting on the afferent terminals to improve vesicle recovery (hypothetically by disabling inhibition of calcium influx from terminal GABAB receptors and facilitating vesicle recycling^43,65^, H3 Figure 4A). If this were the case, peak-to-peak response amplitudes would be higher, post-activation depression would be reduced due to the greater pool of available vesicles, and there should consequently be a slowed rate of decay (H3). In summary, no change in the decay rate would indicate that the CST is only acting post-synaptically to depolarize motoneurons (H1), a faster decay would indicate that volitional input is facilitating reflex responses via PAD (H2), and a slower decay would suggest that volitional input is facilitating reflex responses at the presynaptic terminal (H3). (Figure 4A–B)

To verify the validity of our conjectures we used a simple simulation of motoneuronal activation^33^ (Figure 4C). The model simulated a pool of motoneurons receiving excitatory input from damaged supraspinal fibers and SCS-mediated sensory afferents, both of which were treated as a probabilistic triggering of EPSPs. From the motoneuron pool firing times, we derived an estimated EMG signal. To incorporate CST effects on presynaptic post-activation depression in the afferents, we integrated a previously-validated probabilistic model of spike transmission failure which governed the amplitude of the SCS-mediated EPSPs^66^. In this model, there are two potential points of spike transmission failure from SCS to the motoneuron. The first point occurs in afferent axons, where spikes may fail to propagate (biophysically, this would occur at axonal branchpoints, for which the probability of failure has been shown to be modulated by PAD^42,43^). The second point occurs at the afferent presynaptic terminal, where vesicle depletion can reduce the probability of neurotransmitter release (Figure 4C, see methods).

We then simulated the three different hypotheses: Under H1, only the magnitude of volitional drive (firing rate of the CST) was varied, while presynaptic parameters were held constant across conditions. Under H2, the probability of axonal spike failure (failure point 1, Figure 4C) was decreased proportionally with the magnitude of volitional drive, reflecting a PAD-like decrease in spike failure at the afferent branch points. Lastly, under H3, the rate of vesicle recovery was increased proportionally to the magnitude of volitional drive, producing faster recovery and a reduced probability of synaptic transmission failure (failure point 2, Figure 4C).

Across all model hypotheses, simulated responses decayed as an exponential across time. In the simulated H1 condition, an increase in CST drive led to stronger reflexes, but no change in the exponential decay time constant (τ_1_, Figure 4D–G). Under H2, decreased probability of axonal failure also led to stronger reflex responses (Figure 4H–I), but the decay time constants quickened with increasing CST drive (Figure 4I–K). Lastly, under H3, we again saw stronger reflexes(Figure 4L–M), but decay time constants slowed with increasing CST drive(Figure 4M–O). Therefore, this model indicates that our prediction is plausible and that we can use the change in experimental rates of decay of SCS trains to distinguish between post-synaptic potentiation of motoneuron excitability (H1) and pre-synaptic gating of afferent transmission (H2–H3).

We tested these hypotheses with in-vivo experiments. Participants were asked to rest or maintain a constant force of either 10% or 25% of their MVC during elbow extension or flexion. This produced three different supraspinal input levels. During the force hold, 2s bursts of stimulation were applied at 5hz, 40hz, and 80hz. Importantly, post-activation depression exhibits a frequency dependent effect: at higher stimulation rates of the dorsal roots, the attenuation of responses progressively increases^23,38,67^. An example of this is shown in Figure 5A. When comparing the average response across the stimulation burst, it is clear that there are smaller responses at higher frequencies (Figure 5B). As such, we would expect to see the strongest attenuation of SCS-evoked responses when SCS is delivered at 40 or 80hz. Therefore we focused the subsequent analysis on 80hz stimulation trains.

**Figure 5.**
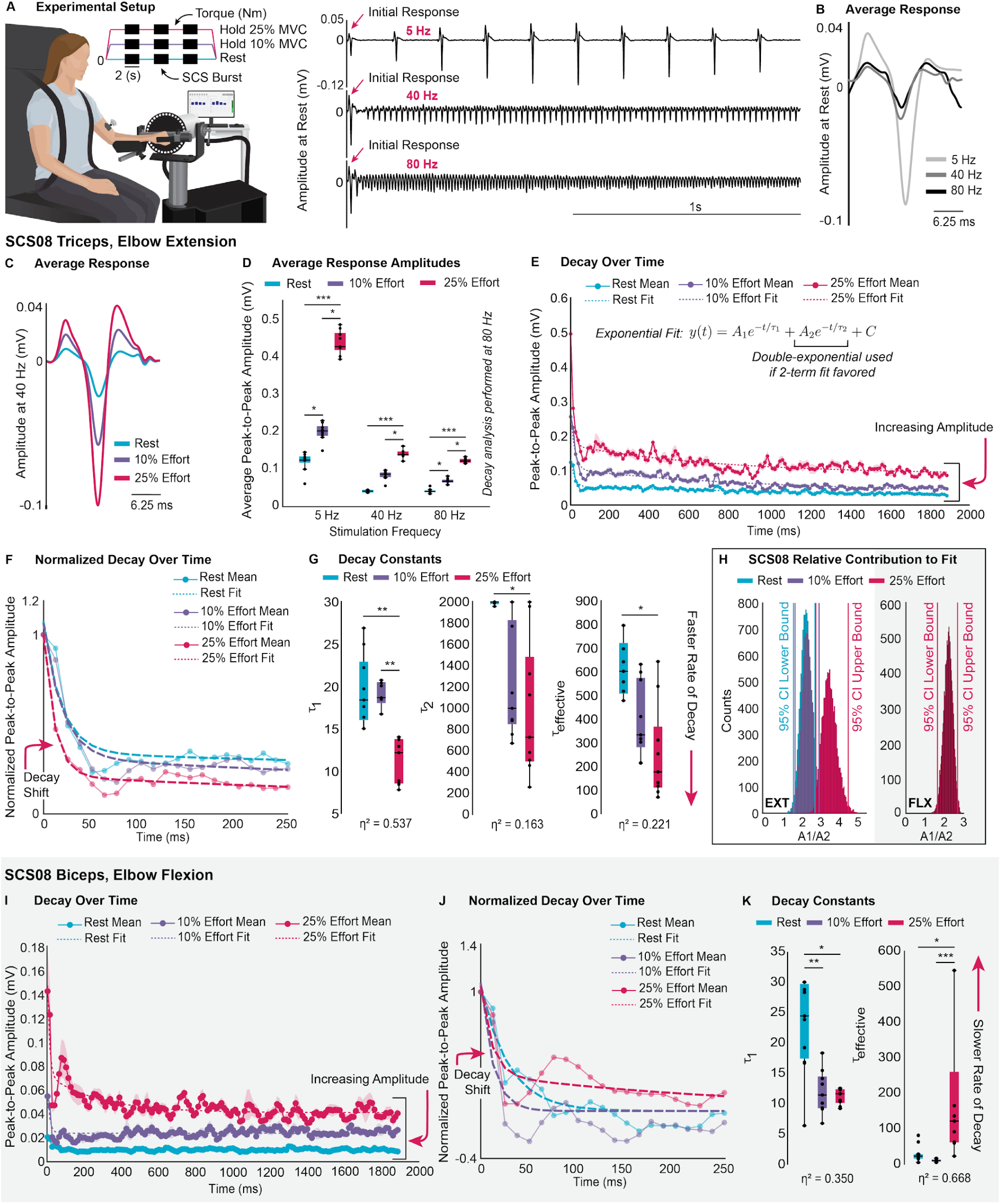
In vivo: volitional modulation of post-activation depression demonstrates presynaptic control of sensory inputs. **(A)** Left: experimental setup. Right: example SCS-evoked responses at different frequencies at rest demonstrating rate-dependent post-activation depression. **(B)** Example average waveform at rest across frequencies (light grey: 5hz; medium grey: 40hz; black: 80hz) **(C–G)** SCS08, elbow extension, triceps. **(C)** Average waveform at 40hz across effort levels. **(D)** Boxplots (median, IQR, whiskers = 1.5×IQR) showing mean peak-to-peak amplitudes for each burst across frequencies and effort levels with overlaid dots (n=9 bursts). **(E)** Peak-to-peak amplitude across 80hz bursts (mean (solid line) ± SEM; double exponential fit (dashed line); n=9 bursts). **(F)** Normalized peak-to-peak amplitude across 80hz burst (mean (solid line); double exponential fit (dashed line); n=9 bursts). **(G)** Boxplots (median, IQR, whiskers = 1.5×IQR) summarize fitted decay time constants (τ_1_, τ_2_, τ_*eff*_) with overlaid dots (n=9 bursts). **(H)** Relative contribution of first and second exponential decay components (*A*_1_ /*A*_2_). Bootstrapped 95% CIs of the mean (10,000 resamples) shown in corresponding colors. Left: SCS08, extension, triceps. Right: SCS08, flexion, biceps. Ratios > 1 indicate τ_1_ as the primary decay constant. **(I–K)** SCS08, elbow flexion, biceps. **(I)** Peak-to-peak amplitude across 80hz burst (mean (solid line) ± SEM; exponential fit (dashed line); double-exponential used if two-term model favored; n=9 bursts). **(J)** Normalized peak-to-peak amplitude across 80hz burst (mean (solid line); exponential fit (dashed line); double-exponential used if two-term model favored; n=9 bursts). **(K)** Boxplots (median, IQR, whiskers = 1.5×IQR) summarize fitted decay time constants (τ_1_, τ_*eff*_) with overlaid dots (n=9 bursts). *Rest: blue; 10% effort: purple; 25% effort: pink. Panels D, G, K: Kruskal-Wallis with Tukey-Kramer post hoc, *p<0*.*05, **p<0*.*01, ***p<0*.*001. Effect size reported as η*^*2*^.

First, as predicted by the model, the amplitude of SCS-evoked responses increased with increasing effort (Figure 5C–D) and showed measurable decay over time. In participant SCS08 during elbow extension, the decay of SCS-evoked responses was well-fit by a double-exponential function. Increasing effort produced clear increases in response amplitudes across the bursts of stimulation (Figure 5E) as well as a distinct acceleration in decay (Figure 5F). Fitted decay time constants (τ_1_, τ_2_) decreased systematically with effort, resulting in a faster effective rate of decay (Median τ_*eff*_=600.3ms at rest, 330.8ms at 10%, and 17.4ms at 25%; Figure 5G). The relative weighting of the fast and slow components (Figure 5H, left plot) showed that the rapid, early decay—captured by τ_1_ (Figure 5F)—was the dominant contributor to this change. A similar trend was observed in participant SCS07 during elbow flexion. As in SCS08, response amplitudes increased and decay time decreased with effort (Median τ_1_=18.05ms at rest, 29.91ms at 10%, and 5ms at 25%; Figure S3A–C). These findings are strongly consistent with H2, in which residual CST inputs act presynaptically via PAD to depolarize sensory afferents.

Interestingly, evidence from SCS08 during elbow flexion suggests that the CST may also modulate sensory afferent transmission via the mechanism proposed in H3. At rest and 10% effort, the decay was best fit by a single-exponential function, whereas at 25% effort a double-exponential model provided the best fit. As in extension, response amplitudes increased with effort, however, the early portion of the decay was visibly faster with increasing drive (Figure 5I–J). The fitted decay time constants confirmed that τ_1_ decreased with effort (Median τ_1_=24.57ms at rest, 11.54ms at 10%, and 11.74ms at 25%; Figure 5K). However, calculation of τ_*eff*_ revealed significantly slower decay at 25% effort (Median τ_*eff*_=24.57ms at rest, 11.54ms at 10%, and 123.2ms at 25%). While the initial drop in amplitude (captured by τ_1_) remains the dominant component of post-activation depression (Figure 5H, right plot), this finding points to an additional slower component of recovery that can be engaged by supraspinal connections, consistent with the modulatory effect predicted by H3.

Another case comes from SCS07 during elbow extension (Supplemental Figure S3D–F), where the decay dynamics remained consistent across effort levels. Although response amplitudes increased with 25% effort (Figure S3D), neither τ_1_ nor τ_*eff*_ differed significantly with effort (Figure S3E–F). This suggests that in some instances facilitation may arise predominantly through postsynaptic mechanisms, consistent with H1, wherein CST drive enhances motoneuron excitability without visibly altering presynaptic dynamics.

Taken together, these results demonstrate that residual supraspinal connections can shape the excitatory drive of SCS to motoneurons; predominantly by facilitating Ia-afferents (H2), but potentially also by enhancing vesicle recovery to lessen post-activation depression (H3). Thus, we show that supraspinal input presynaptically modulates SCS-evoked reflexes, likely through CST-polysynaptic mechanisms that regulate sensory afferent transmission.

### Supraspinal drive sculpts SCS-engaged stretch reflexes during functional movement

We showed that residual CST projections can modulate reflexes via presynaptic mechanisms; but what is the relevance of these mechanisms during functional movement? To assess this, we developed a protocol in which test pulses of SCS were delivered to evoke clear reflex responses during movement (Figure 6A). Because the stretch reflex, a spinal circuit in which muscle spindle afferents excite motoneurons of the same muscle, engages the same afferent-motoneuron pathway as SCS, we expected SCS-evoked responses to be modulated by joint-angle over time: e.g. reflexes should increase in amplitude as the muscle lengthens and spindle input rises, and decrease as the muscle shortens^68,69^ (Figure 6B). This relationship is exemplified in Figure 6A during passive movement and 10hz stimulation of the biceps. By quantifying this stretch-reflex–like pattern of SCS activation (amplitude-angle slope relationship) during passive and active movement, we could directly assess the innate capacity of the residual CST to modulate spinal reflexes during dynamic movements.

**Figure 6.**
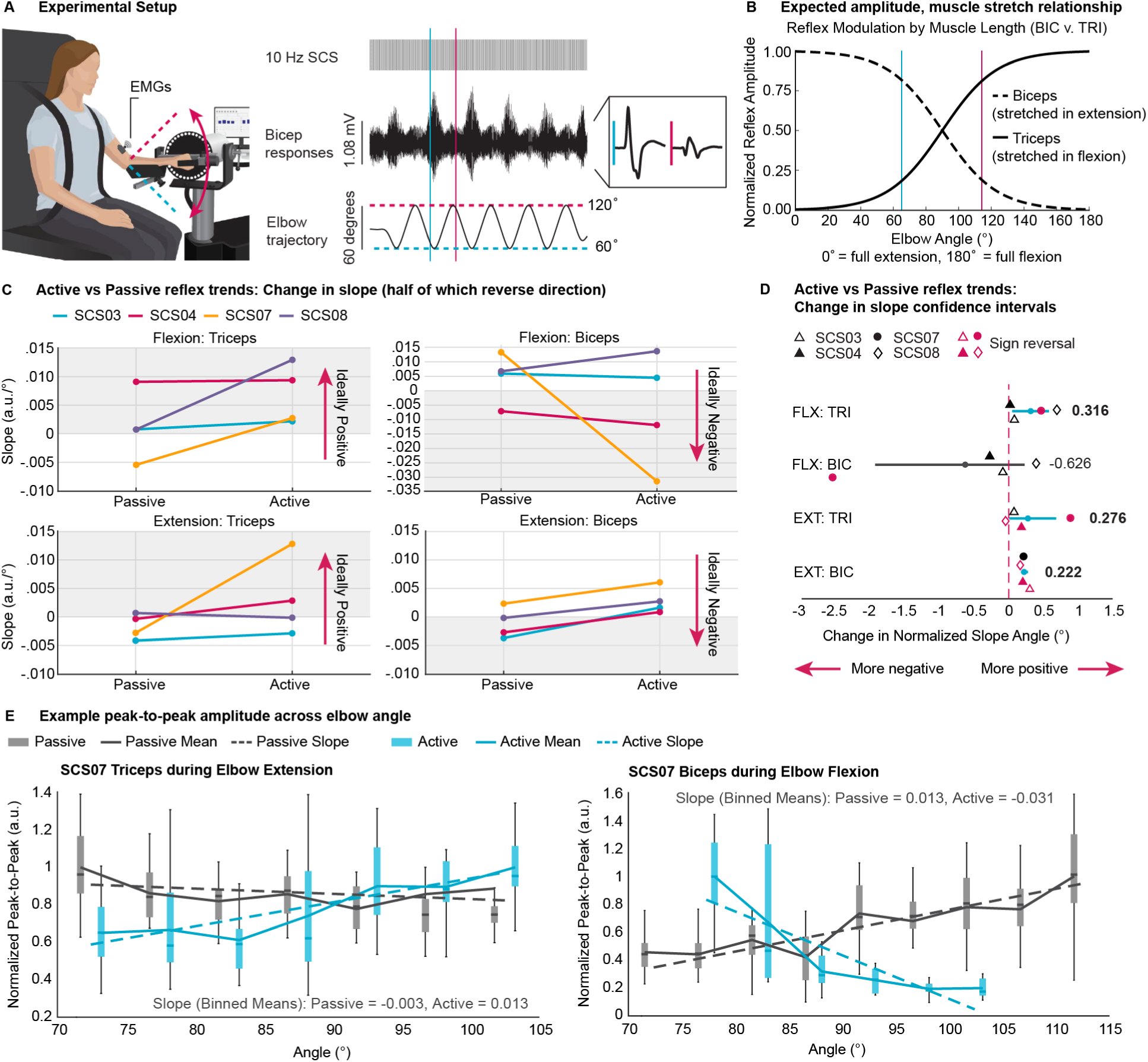
Supraspinal drive sculpts SCS-engaged stretch reflexes during functional movement. **(A)** Experimental setup. **(B)** SCS recruits the sensory-motor reflex arc on top of the intrinsic muscle stretch reflex, which follows a well-defined amplitude-angle relationship. Shown is the expected relationship between biceps (dashed line) and triceps (solid line) across elbow angle. Reflex amplitude increases as each muscle is stretched. Vertical lines in blue and pink correspond to the example time points in (A), where this relationship is observed in biceps. **(C)** Participant-level slopes (normalized by the peak mean amplitude across bins) showing differences in reflex amplitude-angle relationship during passive and active conditions. Shaded regions indicate expected slope sign (Biceps –; Triceps +). Lines which cross from unshaded to shaded, or vice-versa, indicate a reversal of amplitude-angle relationship with supraspinal input. **(D)** Group-level changes in normalized slope angle (delta degree) between active and passive conditions. *Participant-level deltas resampled with replacement (10,000 iterations) to estimate the group-level mean and 95% CI. CIs excluding zero (blue) indicate significant changes; mean change in degrees shown for each interval*. **(E)** Example normalized peak-to-peak amplitudes binned across elbow angle from SCS07 (boxplots: median, IQR, whiskers = 1.5×IQR). Left: triceps during extension; right: biceps during flexion. Passive movement (grey); active movement (blue). Solid lines show mean values; dashed lines show linear fits. Reported slopes correspond to fits of binned means. Both show strong reversal of amplitude-angle relationship.

Interestingly, during passive movement, the pattern of reflex modulation revealed evidence for disrupted spinal circuitry after stroke. While some participants followed the expected stretch-reflex–like pattern wherein responses increased as the muscle lengthened, half of the recorded passive conditions across the biceps and triceps showed inverted relationships in which reflex amplitudes decreased during stretch (Figure 6C). This inverted slope likely reflects a breakdown in normal coordination between agonist and antagonist muscles, consistent with impaired reciprocal inhibition^70–73^. Such pathology further distorts the baseline sensorimotor reflexes on which volitional descending input must act to regain coordinated movement.

With the introduction of volitional drive, however, many of these abnormal amplitude-angle relationships observed during passive movement reorganized, even inverted, towards the expected physiological stretch-reflex–like pattern. Indeed, we found across participants, the amplitude-angle relationship changed both significantly and appropriately between active and passive conditions, as quantified by the change in normalized slope angle (°). This effect was particularly evident for triceps during both elbow flexion (mean change: +0.316, CI [0.049, 0.584]) and extension (mean change: +0.276, CI: [0.011, 0.690]) (Figure 6C-D). Particularly striking was SCS07 wherein both the triceps and biceps inverted to the expected orientation during active movement (Figure 6E), demonstrating a clear capacity to sculpt reflexes during movement. It should be noted however, that bicep responses were more heterogeneous; some moved toward the expected negative orientation, others counterintuitively became more positive. However, given the canonical post-stroke flexor bias, where flexor muscles (e.g., biceps) are over-excited^7^, this is somewhat unsurprising. These findings demonstrate that the polysynaptic CST-dependent capacity to sculpt sensory-motor reflexes (e.g. SCS, stretch-reflex) is used during movement and can normalize pathological spinal circuit behavior.

### Supraspinal sculpting of SCS reflexes is strongest during CST-dominant motor control

Up until this point, we have primarily used SCS to evoke reflex responses and probe intrinsic properties of cortico-spinal circuitry. And while we have shown that SCS can be used to improve gross and fine motor function (Figure 1), is residual supraspinal input, from the CST in particular, capable of modifying the effects of SCS at therapeutic settings to produce functional muscle activations? To test this, we directly compared motor performance and muscle activation patterns between the previously described MVC and force control tasks (20–35% MVC) (Figure 7A). This design allows for direct comparisons between conditions that are executed with the same isometric setup and SCS parameters, differing only in volitional demand. In particular, fine force control forces participants to use their residual CST connections to regulate muscle activity. Contrastingly, MVC favors less refined control policies such as excitation from the RST^6,11,74–76^.

**Figure 7.**
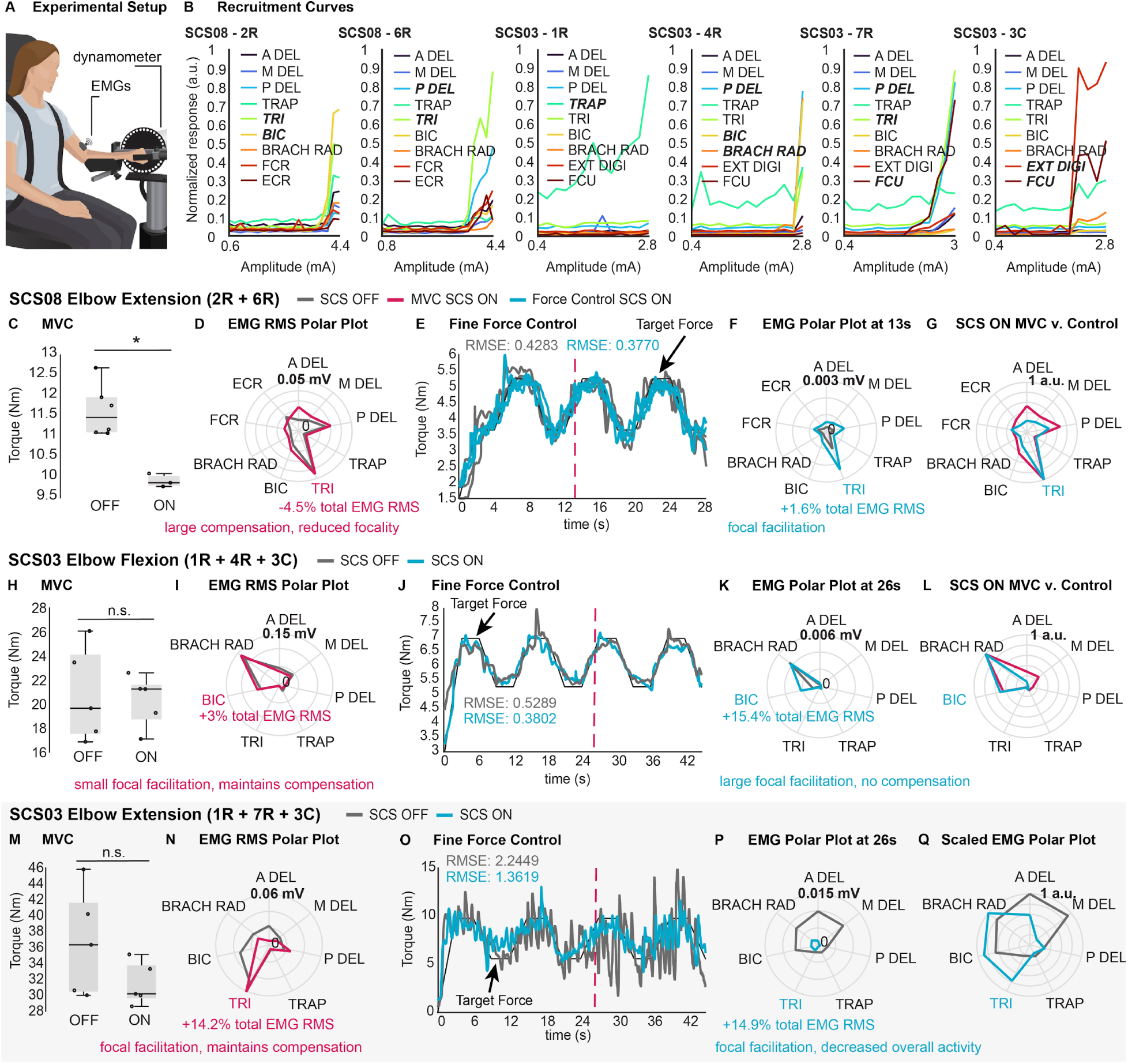
Supraspinal sculpting of SCS reflexes is strongest during CST-dominant motor control. **(A)**Experimental setup. The same setup and stimulation configurations were used in the shown MVC and corresponding fine-force control tasks. **(B)** Recruitment curves (response amplitude by stimulation amplitude). Each panel corresponds to a unique stimulation contact in relevant participants. Colors denote individual muscles. **(C–G)** SCS08 elbow extension; stimulation contacts 2R+6R. **(C)** Boxplots (median, IQR, whiskers = 1.5×IQR) showing MVC torque for SCS OFF vs. SCS ON with overlaid dots (n=3–6 per condition). **(D)** Polar plots of EMG RMS activity. Each radial line represents a recorded muscle during MVC. SCS OFF (grey); SCS ON (pink). The primary agonist muscle and the percent change in primary agonist to total EMG RMS are indicated in pink. **(E)** Mean fine-force control torque traces showing decreased error (RMSE) during SCS OFF (grey) vs. SCS ON (blue). Individual repetitions shown with reduced opacity (n=3 per condition). **(F)** Polar plot of EMG RMS activity at indicated time-point. Each radial line represents a recorded muscle during force-tracing. SCS OFF (grey); SCS ON (blue). The primary agonist muscle and the percent change in primary agonist to total EMG RMS are indicated in blue. **(G)** Polar plot of EMG RMS activity during SCS ON compared between MVC (pink) and fine force control (blue). Each radial line represents a recorded muscle normalized to the muscle with maximal activation during MVC or fine force control respectively. **(H–L)** SCS03 elbow flexion; stimulation contacts 1R+4R+3C. Same format as **(C–G)**. MVC n=5 per condition; fine force control n=1 per condition. **(M–Q)** SCS03 elbow extension; stimulation contacts 1R+7R+3C. (M-P) Same format as **(C–F)**. MVC n=5 per condition; fine force control n=1 per condition. **(Q)** Scaled polar plot of EMG RMS activity from (P). Each radial line represents a recorded muscle normalized to the muscle with maximal activation during SCS OFF (grey) or SCS ON (blue) respectively. *Panels C, H, M: Inference on mean differences in MVC was performed using bootstrap resampling (n=3–6 repetitions per condition, 10,000 samples). Asterisks denote statistical significance, defined as rejection of the null hypothesis of no difference at the 95% confidence interval. A DEL: anterior deltoid, M DEL: medial deltoid, P DEL: posterior deltoid, TRAP: trapezius, TRI: tricep, BIC: bicep, BRACHRAD: brachioradialis, EXTDIGI: extensor digitorum, ECR: extensor carpi radialis, FCR: flexor carpi radialis, FCU: flexor carpi ulnaris*.

Importantly, we observed that SCS sometimes failed to improve or even worsened performance during MVC, yet would still enhance accuracy during force control. It is these examples that we dissected. For example, in SCS08 during elbow extension, SCS reduced MVC torque while broadly facilitating activity across multiple muscles, decreasing the relative contribution of the agonist triceps (−4.5% of total RMS across muscles; Figure 7C–D). This reduction suggests that non-specific supraspinal drive, coupled with the limited selectivity of SCS, can result in the concurrent amplification of antagonist or compensatory muscles, thereby hindering functional torque production. In contrast, during force control, accuracy improved by 11.9% and was accompanied by a more selective triceps activation with SCS (+1.6% of total RMS across muscles; Figure 7E–F). Direct comparison of muscle activation patterns between tasks in the SCS on condition further reveals a reduction of compensatory activity during fine force control (Figure 7G). Thus, both performance and the underlying muscle activation profile differ markedly between MVC and force control tasks. Similar task-dependent changes were observed in SCS03 during elbow flexion (Figure 7H–L).

Interestingly, during elbow extension in SCS03, improvements in force control accuracy arise in a seemingly different manner. Rather than broad facilitation with task-dependent differences in selectivity, SCS instead produced an overall reduction in muscle activity during fine force control, accompanied by a pronounced redistribution toward the agonist triceps (+14.9% of total RMS across muscles; Figure 7P–Q). In contrast, during the MVC task, facilitation of both agonist and compensatory muscles was retained; therefore, even when activation appeared more focal, there was no corresponding improvement in maximum torque (Figure 7M–N).

Together, these examples demonstrate that when a non-specific or aberrant supraspinal input is used to activate muscles (as during MVC in our stroke participants), SCS can become ineffective and potentially detrimental. This is because dorsal roots project to multiple muscles, including antagonists; as a result, SCS is poorly selective and can amplify nonfunctional muscle activity, effectively worsening an already “incorrect” motor command. In contrast, when participants performed tasks that explicitly required engagement of residual CST pathways, they were capable of steering the excitatory effect of SCS towards the relevant muscles, consistent with our findings. This demonstrates that residual CST activity can sculpt SCS-mediated muscle activity for skilled force control, and that improvements in fine motor performance arose not simply from the addition of SCS, but from the engagement of a CST-oriented motor control strategy.

## DISCUSSION

We showed that people with stroke can partially recover arm dexterity through poly-synaptic CST pathways, which primarily involve pre-synaptic modulation of primary afferents. Our results challenge the assumption that fine motor control of the human arm and hand is solely dependent on monosynaptic CST connections to the motoneurons.

### Polysynaptic circuits involved in the recovery of fine motor function with neurostimulation

Our results point to at least two independent polysynaptic pathways that could be involved in the recovery of motoneuron control post-stroke. The first pathway would act via post-synaptic modulation of motoneuron excitability and a second pathway would act via pre-synaptic modulation of the primary afferents.

First, we found substantial facilitation of spinal reflexes by TMS conditioning at latencies compatible with one or few excitatory synapses between the cortico-motoneuron axon and the spinal motoneurons which supports the existence of excitatory polysynaptic CST connections to motoneurons, particularly given the reverse protocol showed no facilitation even with short ISIs.

Second, TMS pulses produced strong reflex facilitation at long ISIs (80–300ms). These timings are strikingly similar to peak facilitatory action caused by PAD^42^, a GABAa receptor-mediated facilitation of primary afferent conduction^43,44^. Indeed, single-motoneuron discharges in response to TMS showed that post-synaptic facilitation of motoneurons was only present at short ISIs. Therefore, since no change, and even reductions in motoneuron firing rate occurred at intervals longer than 80ms, the strong reflex facilitation at long ISIs can only be explained by a pre-synaptic potentiation of primary afferent inputs to motoneurons. Furthermore, the modification of spinal reflex decay rate during post-activation depression is consistent with H2, e.g. that the CST facilitates afferent activity and neurotransmitter release via an axo-axonic mechanism (like PAD).

Together with evidence of reflex modulation and inhibition we showed in our experiments, our findings demonstrate that residual CST axons can finely tune spinal circuit excitability through a variety of polysynaptic mechanisms, the most consistent and prominent of which seems to be presynaptic modulation of sensory transmission.

### Relevance of polysynaptic cortico-spinal pathways for motor control

These reflex-modulating capabilities were not dependent on the presence of SCS. Rather, our participants were able to modulate evoked sensorimotor reflexes without assistive SCS (Figure 6); but what is the relevance of these intrinsic capabilities for dexterous motor control?

Motor control requires both the ability to generate excitatory drive to activate spinal motoneurons and the ability to direct that excitation toward functionally relevant muscles. The monosynaptic CST provides excitation with high selectivity, thereby enabling dexterity and single digit control^1–6^. In contrast, polysynaptic excitatory pathways to motoneurons^2,3,5,16–19^ are considered less selective due to divergent connectivity^77^. Indeed, after stroke, our participants retained the capacity to activate muscles, but displayed substantial co-contraction, consistent with reduced selectivity.

Our results reveal, however, that corticospinal fine motor control could be exerted, at least in part, through polysynaptic descending pathways responsible for presynaptic modulation of primary afferents^1–3,42,78,79^. Indeed, spinal excitability can be functionally tuned pre-synaptically with spatial and temporal precision^44,80–82^. Consistent with this, our participants could engage these pre-synaptic pathways during functional movement to sculpt reflex excitability, underscoring a role in motor control. However, their use was imperfect; while sensorimotor reflex patterns were modulated appropriately in some muscles, and even improved, others, such as the biceps, showed exacerbated physiological stroke phenotypes when comparing passive vs. active elbow stretch (Figure 6). This indicates that these polysynaptic pathways, while functional, are still impaired post-stroke^7,70–73^.

Within this context, residual polysynaptic mechanisms likely retain some functionality and can be actively used to shape reflex excitability during movement. This engagement highlights a preserved capacity for volitional sculpting of spinal circuits, even after severe corticospinal damage. However, because these pathways cannot both precisely steer and independently generate excitatory drive, their contribution alone is not sufficient to fully restore dexterous motor control.

### Relevance of polysynaptic cortico-spinal pathways for motor recovery

Analysis of muscle activation underlying the improvements of dexterous force control demonstrate the efficacy of these residual polysynaptic pathways when combined with therapeutic SCS. We showed examples in which our study participants attempted to produce a MVC; an action during which all available excitatory drive is used. It’s believed that the RST takes over some of the excitatory CST function in stroke which leads to unspecific muscle activations^83,84^. Indeed, during these MVC experiments, this strong unspecific excitatory drive (as shown to be present in the SCS off condition), combined with the broadly excitatory effect of SCS, produced large simultaneous potentiation of agonist, antagonist, and compensatory muscles. This not only failed to improve torque, but even impaired it in some cases. This occurred because the unspecific and unsculpted SCS was potentiated by an unspecific supraspinal drive. Instead, when we forced our participants to use their CST to finely regulate force during the same movement, the muscles potentiated by SCS were substantially different, and often reduced in strength to improve torque smoothness, demonstrating an ability to functionally steer and modulate SCS excitatory drive.

This distinction aligns with the broader functional divide between descending systems: gross strength tasks are largely supported by the RST that promote diffuse, synergistic activation, whereas dexterous tasks depend on CST projections capable of selectively shaping motor output. Accordingly, SCS may be most effective when paired with tasks that leverage polysynaptic CST pathways, revealing an inherent and potentially trainable capacity for stroke-affected individuals to reestablish selective control. These results lead to two clinically relevant conclusions, first, MEP status should not be a determinant of who can be trained to use their residual CST capacities because all these abilities that we show occurred in people that were overwhelmingly MEP negative. Indeed, recent work supports separation of MEP status from recovery prognostics^85–87^. Second, physical therapy interventions, in combination with SCS, should focus on promoting CST-skewed exercises rather than a gross strength-producing oriented function in order to maximally benefit from the effects of SCS.

### Limitations of the study

We cannot exclude that some participants retained silent monosynaptic connections to spinal motoneurons. However, modulation of spinal reflexes was reproduced even in individuals with very severe lesions that likely destroyed functional mono-synaptic connections, suggesting that polysynaptic effects are a strong component of the effects we observed. Furthermore, strength gains have previously been shown to converge across MEP-positive and MEP-negative participants post-stroke^86^. Likewise, improvements within the Action-Research-Arm-Test and related functional measures have been reported despite no change in TMS stimulus-response recruitment curves over a rehabilitation period^87^, indicating that behavioral recovery need not mirror strengthened monosynaptic CST transmission. Furthermore, although beta-band corticomuscular coherence is primarily attributed to fast corticospinal fibers, beta-band activity may still travel through polysynaptic pathways, particularly after injury^49^. Critically, changes in coherence may arise from many sources including enhanced motor-unit recruitment or engagement of indirect circuits. When one considers the preservation of dexterous proximal control in species lacking direct cortico-motoneuronal projections^88^, prior clinical and physiological findings in stroke^85–87^, together with our results, a broad consilience emerges supporting a substantial role for recovery of dexterous motor control via non-monosynaptic corticospinal pathways.

While these limitations hinder our ability to definitively conclude that dexterity can be produced by CST poly-synaptic pathways in able bodied individuals, our results demonstrate that it is possible to regain such abilities post-stroke by leveraging the residual CST polysynaptic system.

## Data Availability

All data produced in the present study are available upon reasonable request to the authors.

## RESOURCE AVAILABILITY

### Lead contact

Requests for further information and resources should be directed to lead contact Marco Capogrosso.

### Materials availability

This study did not generate new materials.

### Data and code availability

The main data supporting the results in this study are available within the paper. Raw data will be available upon reasonable request to the corresponding author. Data from the corresponding clinical trial is available at Dabi https://dabi.loni.usc.edu/dsi/UG3NS123135. The code for performing the neural simulations is available at GitHub https://github.com/jostrows9/NonMonosynapticRecoveryPostStrokeModel. Any additional information required to reanalyze the data reported in this work is available from the lead contact upon request.

## ACKNOWLEDGMENTS

The study was executed through the support of National Institutes of Health Brain Initiative grant no. UG3NS123135-01A1 to M.C. and D.J.W. and internal funding from the Department of Neurological Surgery at the University of Pittsburgh to M.C., the Department of Mechanical Engineering and the Neuroscience Institute at Carnegie Mellon University to D.J.W. and the Department of Physical Medicine and Rehabilitation at the University of Pittsburgh to E.P.

## AUTHOR CONTRIBUTIONS

E.S. and M.C. conceived the study. E.S., M.C., J.W.K. and M.G. designed the experiments. M.C., L.E.F. and P.G. designed the neurosurgical approach. G.F.W. and R.M.dF. implemented patient recruitment, eligibility, monitoring and coordinated management of the study. P.G. performed the surgery. E.S., L.B., R.M.dF., and N.V. performed the experiments. J.O. and M.C. developed and performed computational simulations. E.S., L.B., and J.O. performed data analyses and prepared figures. E.S. and M.C. wrote the original draft. All the authors contributed to its review and editing. M.C., D.J.W. and E.P. secured the funding. M.C. and J.W.K. supervised the work.

## DECLARATION OF INTERESTS

D.J.W., P.G., and M.C. are founders and shareholders of Reach Neuro, a company developing SCS technologies for stroke. E.P. has interest in Reach Neuro due to personal relationships with M.C.

E.S., N.V., D.J.W., L.E.F., P.G., E.P., and M.C. are inventors on patents related to this work. All other authors declare no competing interests.

## DECLARATION OF GENERATIVE AI AND AI-ASSISTED TECHNOLOGIES

During the preparation of this work, authors occasionally used ChatGPT (OpenAI, GPT-5) to refine sentence structure for clarity and conciseness. After using this tool or service, authors reviewed and edited the content and take full responsibility for the content of the publication.

## METHODS

### Trials information

All experimental protocols were approved by the University of Pittsburgh Institutional Review Board (IRB) (protocol STUDY19090210) under an abbreviated investigational device exemption. The study protocol is published on ClinicalTrials.gov(NCT04512690). Eight human participants were enrolled in the study, however, only seven completed the study. Specifically, SCS06 was withdrawn from the trial after being diagnosed with arrhythmia during preoperative assessments. All participants provided informed written consent according to the procedure approved by the IRB of the University of Pittsburgh and participants were compensated for each day of the trial and for travel and lodging during the study period. Detailed information including study design, inclusion criteria, and primary/secondary trial outcomes can be found at ClinicalTrials.gov and in the two publications reporting directly on the pilot trial^23,24^.

### Study participants information

SCS01 is a female who had a right hemorrhagic stroke secondary to a thalamic/midbrain cavernous malformation nine years before participating in the study. At the time of the study, she had left-sided spastic hemiparesis. Her spasticity was previously managed with botulinum neurotoxin injections to the biceps, brachioradialis, and pronator teres. These injections were suspended six months prior to and throughout the study. She had also undergone flexor tendon lengthening surgery and a C5–C6 anterior cervical discectomy and fusion in the interim.

SCS02 is a female who had a right ischemic MCA stroke secondary to a right carotid dissection resulting in a large MCA territory infarct 3 years before participating in the study. Her post-stroke residual at the time of participation was a left-sided spastic hemiparesis complicated by a left wrist flexion contracture despite treatment with splinting.

SCS03 is a male who had a hemorrhagic stroke in the right MCA territory, followed by craniotomy and hematoma evacuation seven years prior to the study. At the time of study participation, he had left-sided spastic hemiparesis. He was not taking medications for spasticity but used a home shockwave device for spasticity management and actively attended outpatient physical therapy three times per week.

SCS04 is a male who had an ischemic stroke in the right MCA territory five years prior to study participation. He presented with left-sided hemiparesis and underwent thrombolysis. He completed both inpatient and outpatient rehabilitation and was not being managed for his spasticity at the time of the study.

SCS05 is a male who had a left basal ganglia intraparenchymal hemorrhage two years prior to study participation, resulting in right-sided hemiparesis and transient speech difficulties. MRI confirmed a basal ganglia hemorrhage without underlying vascular abnormalities, consistent with a likely hypertensive etiology. His post-stroke rehabilitation included physical, occupational, and speech therapy with limited functional improvement. He received botulinum neurotoxin injections for spasticity, which were discontinued 10 weeks prior to and throughout the duration of the study.

SCS07 is a female who sustained a left hemispheric lacunar infarct ten years prior to study participation, with a past medical history of hypertension and diabetes mellitus. Her post-stroke deficits included weakness and spasticity in her right upper limb. She actively participated in physical and occupational therapy prior to the study. She also had a joint contracture in the shoulder joint. She weaned off baclofen prior to enrolling in the study.

SCS08 is a female who had a right-sided ischemic stroke involving the corona radiata four years prior to the study. She has a notable past medical history of hypertension. Her post-stroke deficits include persistent weakness and spasticity in the left upper and lower extremities, with preserved sensation in the affected arm. She underwent inpatient rehabilitation following her stroke and was followed by 6 months of at-home physical and occupational therapy. She was managed with continual botulinum neurotoxin injections for left hand and thumb tightness, but these were held for four months prior to and during the trial.

### Intraoperative procedures

Participants underwent percutaneous implantation of two 8-contact leads into the epidural space of the cervical spinal cord. Implantation was performed under general anesthesia with intraoperative neuromonitoring used to guide rostral-caudal placement from C3 to T1, ensuring recruitment coverage from proximal shoulder to intrinsic hand muscles. During the procedure, stimulation selectivity and correct lead placement were confirmed by generating recruitment curves at each contact using a monopolar intraoperative neuromonitoring system (Xltek Protektor, Natus Medical) with a subdermal return electrode placed contralateral to the affected arm. Stimulation pulses were delivered at 1-2 Hz while current was ramped from 0 to 5 mA. Compound muscle action potentials (CMAPs) were recorded bilaterally using subdermal needle electrodes across a broad set of muscles (including trapezius, deltoid heads, biceps, triceps, forearm flexors/extensors, and intrinsic hand muscles). Contralateral recordings ensured that SCS did not induce cross-over effects. After securing the leads, stimulation specificity was further evaluated using frequency-dependent suppression to confirm dorsal afferent activation rather than direct motor efferent recruitment. At study completion, the leads were removed. Full procedural details, including follow-up x-rays to ensure stable lead placement, can be found in the two publications reporting directly on the pilot trial^23,24^.

### Stimulation calibration

SCS was delivered using a clinical-grade current-controlled stimulator (DS8R, Digitimer) paired with a high-compliance 1×8 multiplexer and a custom microcontroller-based control unit that set pulse timing, amplitude, and electrode channel. The system generated cathodic-first, charge-balanced biphasic pulses (200-400 μs) and enabled semi-synchronous stimulation across multiple contacts by rapidly switching channels between pulses. All stimulation parameters including frequency, channel selection, duration, latency, and amplitude were configured through a custom MATLAB GUI communicating with the controller via a defined command protocol. Full technical details can be found in the two publications reporting directly on the pilot trial^23,24^.

#### Recruitment curves

To evaluate the selectivity of SCS in recruiting individual muscles, recruitment curves were generated by measuring the peak-to-peak amplitude of CMAPs as a function of stimulation amplitude. We used a wireless EMG system (Delsys, Trigno) to record the CMAPs from multiple upper-limb muscles, such as trapezius, anterior deltoids, biceps, triceps, brachioradialis, and abductor pollicis brevis. Stimulation was delivered to one contact at a time at 1-2 Hz, Stimulation pulses were delivered at 1-2 Hz while current was ramped from 0 to 8 mA, or until the participant reported discomfort. Monopolar stimulation was applied using a return surface electrode placed over the iliac crest contralateral to the affected arm. For each muscle, the peak-to-peak amplitude of CMAPs elicited at each stimulation amplitude was normalized to the maximum amplitude obtained across each measured trial. This process was repeated for every contact on both leads, thereby obtaining recruitment curves that described a relationship between SCS amplitude and muscle recruitment.

#### Systematic procedure for SCS calibration

Stimulation parameters were calibrated systematically to determine a subset of contacts which would facilitate voluntary movements across multiple upper-limb joints. First, recruitment curves were used to identify contacts that selectively activated different muscle groups (e.g., elbow flexors vs. extensors). Then, for each selected contact, stimulation amplitude and frequency were individually tuned while participants performed movements involving the joint(s) targeted by stimulation (e.g., elbow flexion vs. extension). Recruitment curves were used as a starting point for determining stimulation amplitude. However, a combination of patient feedback, qualitative assessment, and quantitative performance comparisons were used to finalize the stimulation amplitude (between 0.2 mA and 8 mA) and frequencies (between 40 Hz and 100 Hz) used in behavioral assessments. Pulse-width was typically set to 200 µs. Frequency was only increased when increasing stimulation amplitude failed to produce satisfactory outcomes. After individually calibrating a set of contacts (usually 2 or 3), we combined them with a personalized configuration for each participant. Stimulation amplitudes were re-adjusted when combining multiple contacts to minimize interference between their effects. Less frequently, we also adjusted the stimulation frequency across contacts.

### Maximum voluntary contraction (MVC)

Strength at the elbow was assessed isometrically using a robotic isokinetic machine (HUMAC NORM, CSMi). During testing, the robot’s manipulandum was locked such that the participant’s elbow was flexed to 90° and aligned with the rotation axis of the manipulandum. Participants were then instructed to exert their maximal voluntary force in either flexion or extension for 5 seconds. To ensure isolation of single-joint motion, participants were stabilized using shoulder harnesses, the upper arm and elbow were firmly supported against the chair back, and participants were actively monitored for compensatory strategies. The maximum torque value from each repetition was considered for analysis. Surface electromyography (EMG) was also used to record muscle activity across several muscles of the arm using up to 12 synchronized wireless sensors (Avanti Trigno, Delsys Inc.).

Grip strength was similarly quantified using a hydraulic hand dynamometer (Fabrication Enterprises, no. 12-0021). Participants were instructed to hold the device in a comfortable position and exert maximum grasping force for 5 seconds. The highest value from six repetitions was recorded and used for analysis. Assistive effects of SCS on strength were assessed by comparing MVC force under SCS ON and OFF conditions within the same experimental session.

### Fine force control

Fine force control during elbow flexion and extension was assessed using the same isokinetic machine (HUMAC NORM, CSMi) described above. Participants were instructed to isometrically track a slowly varying target force at two submaximal ranges: 20-35% and 35-50% of their individually measured MVC (assessed with SCS OFF). Counterweights were used where appropriate to offset the mass of the arm and manipulandum, thus allowing for accurate force tracking. Each trial required the participant to meet and follow a predefined force trajectory while the elbow joint remained fixed. The task was repeated under SCS ON and SCS OFF conditions within the same experimental session. The root-mean-square-error (RMSE) of the force trace for each repetition was calculated from the target trajectory and used for analysis. Muscle activity was also recorded during this task using a 64-channel high-density EMG grid (TMSi, The Netherlands) placed over the biceps and triceps, supplemented by surface EMG from other upper-limb muscles using a Delsys sensor configuration identical to that used during MVC testing.

The grasp force-modulation task similarly required participants to produce graded isometric forces at submaximal ranges (20-35% and 35-50% of their maximum grip strength), performed with and without SCS. Grip forces were measured continuously using a 6-axis low-profile force/torque sensor (Mini40, ATI Industrial Automation, North Carolina), allowing precise quantification of force scaling and control.

### Planar reaching

To further assess the assistive effects of SCS on dexterity, we quantified arm kinematics during a planar reach-and-pull task. Specifically, we used a robotic augmented reality exoskeleton system (KINARM, Kingston, ON, Canada). Participants were secured in a wheelchair with adjustable height and limb supports to avoid the contribution of gravity. The task was then displayed onto a dichroic augmented reality display in front of the individuals. Specifically, participants were asked to reach one of the three virtual targets while receiving the feedback of the hand position. On each trial, the task started with the robot moving the participant’s arm to the starting position. To signal the beginning of the movement, an audio cue was played after a randomized time delay of 100-700 ms. The reaching phase was considered completed when the participant kept the index finger for 500 ms within a 0.5 cm radius respect to the target. If participants were unable to reach the target within 10 seconds from the audio cue, the robot moved the arm back to the starting position and the next target was presented. Target position was adjusted with respect to each participant’s range of motion to avoid patient discomfort while performing the task. To quantify movement performance we measured three kinematics metrics for each trial: time to reach the target (s), number of elbow speed peaks (u.a.) and path efficiency (u.a.). The time to reach the target was measured as the time elapsed from the starting audio cue to the completion of the reaching phase. A shorter time indicates faster movements. The smoothness of the movement was quantified as the number of peaks in the elbow speed profile during the same time interval. Fewer peaks indicate smoother movements. The Matlab function *findpeaks* with minimum peak height of 0.1 was used to identify the peaks in the speed profile. Finally, the path efficiency was measured as the Euclidean distance between the starting position and the end target normalized with respect to the total hand path length within the time interval describing the reaching phase. Equation 1 describes the quantification of this metric. A value closer to 0 therefore indicates trajectories closer to a straight line. Data processing and analysis was done with Maltab R2023b (The Mathworks Inc., Natick, MA).

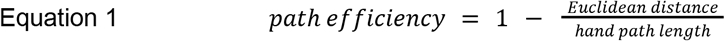

### TMS + SCS paired-pulse protocol

#### TMS and SCS setup

Corticospinal tract integrity was assessed using single pulses of TMS delivered with a Magstim 2002 stimulator and a figure-eight coil. TMS coil placement was guided using stereotactic neuronavigation (BrainSight) co-registered to each participant’s anatomical MRI, with craniofacial landmarks digitized using a Polaris optical tracking system. A 3×3 cm grid centered over the hand knob area of the primary motor cortex in the affected hemisphere was sampled while recording from up to 12 upper-limb muscles using surface EMG electrodes (Avanti Trigno, Delsys Inc.). Participants were classified as MEP+ if peak-to-peak MEP amplitudes ≥50 μV were elicited in any proximal or distal muscle of the impaired arm. The location wherein MEPs were most consistently and strongly elicited was used for later paired-pulse testing. Participants were classified as MEP-if MEPs could not be evoked. For MEP-individuals, hotspot localization was first performed on the contralesional hemisphere. Locations which preferentially activated muscles of the hand were chosen as these tend to consist of dense corticospinal projections. The chosen location was then mirrored to estimate the corresponding location on the lesioned side for later paired-pulse testing. When contralesional stimulation was required for hotspot detection, surface EMG electrodes were also placed on the unimpaired limb.

SCS was delivered through a single contact which corresponded to the same motor pools activated by the chosen TMS hotspot. This was consistently our chosen ‘hand’ contact which selectively recruited distal muscles such as the abductor pollicis brevis, abductor digiti minimi, and the flexor digiti minimi. Recruitment curves for both TMS and SCS were used to verify motor thresholds. Conditioning stimuli were delivered at ~20% below motor threshold and conditioned stimuli at ~20% above threshold. For MEP-participants, the TMS intensity was simply set to 100% of stimulator output.

#### Paired-pulse protocol

Two paired-pulse protocols were performed: SCS-conditioned TMS, in which a single pulse of SCS served as the conditioning stimulus to a single pulse of TMS; and TMS-conditioned SCS, in which a single pulse of SCS served as the the conditioning stimulus to a single pulse of TMS. We employed two AM stimulators (model 2100 A-M Systems, Sequim, WA, USA) to coordinate stimulation delivery. One activated our SCS delivery system while simultaneously sending a pulse to the second AM stimulator which was set to activate the TMS stimulator after a manually defined time interval. Both protocols used the same set of inter-stimulus-intervals (ISIs), spanning from as short as 2ms to as long as 505ms. There was some variability in ISIs used across participants due to differences in stimulation trigger delivery, but manual inspection of EMG stimulation artifacts and recorded trigger pulses ensured accurate interval assignment. For each ISI, a set of approximately 10 repetitions were collected. ISIs were randomized to reduce ordering effects. Furthermore, individual repetitions were separated by ~4s, and sets were separated by ~4min to minimize habituation. The stimulation-triggered response amplitude (of each conditioned stimulus response) was used for statistical analysis. For each recorded muscle, the various recorded peak-to-peak amplitudes were normalized to the mean peak-to-peak amplitude of a given muscle when no conditioning was applied.

To control for auditory effects of TMS which have been shown to co-activate the RST^56^, sham conditioning trials were performed using the same procedure as the TMS-conditioned SCS protocol, but with the coil reversed and held away from the scalp. Only one or two ISIs (a short 10ms interval and a long 100ms interval) were tested in this sham condition.

### Post-synaptic potentials

To estimate the latency and duration of excitatory and inhibitory post-synaptic potentials we performed a cumulative sum (CUSUM) analysis on the peristimulus frequencygrams (PSF)^61^ of the single-motor units detected during a grip force task wherein participants were asked to sustain 20% of their maximum voluntary contraction for 1 minute while holding a handheld dynamometer. Specifically, the CUSUM analysis was performed to identify the point after TMS when a significant change occurred. To assess significance in the deflection of the CUSUM traces we used the error box method. After subtracting the average prestimulus firing rate, we defined the error box as the maximum prestimulus CUSUM deflection, shown as horizontal red lines in the PSF plots. The time point at which the poststimulus CUSUM deflections was larger than the limits of this error box defined the latency of the post-synaptic potentials. An upward significant deflection indicated net excitation (EPSP) and a downward significant deflection indicated net inhibition (IPSP). The time point corresponding to the maximum (EPSP) or minimum (IPSP) poststimulus deflection was then used as measure of the duration of the post-synaptic potentials.

### In-vivo modulation of post-activation depression

To probe presynaptic modulation of SCS-evoked reflexes, a post-activation depression paradigm was used wherein high-frequency stimulation produces characteristic attenuation of successive reflex responses. Participants were seated in the previously described robotic isokinetic machine (HUMAC NORM, CSMi) and positioned for isometric elbow flexion/extension. Three supraspinal drive conditions were tested: rest, steady contraction at 10% of each participant’s MVC, and 25% of each participant’s MVC (assessed with SCS OFF). This was done for both elbow flexion and extension. In all conditions, surface electromyography (EMG) was used to record muscle activity in the agonist muscle (e.g. triceps during elbow extension) (Avanti Trigno, Delsys Inc.). During the steady contraction (or rest), once torque was stable, suprathreshold bursts of SCS were delivered. Each burst lasted 2 seconds and consisted of stimulation at 5, 40, or 80 Hz. Nine bursts were delivered per condition, with at least 2 seconds between bursts. Longer breaks were additionally provided between each set of 3. Stimulation-triggered responses for each pulse of SCS were then extracted for analysis.

The average peak-to-peak amplitude of responses across each burst were used to evaluate differences in potentiation across different frequencies and at different levels of supraspinal drive. At 80 hz, further analysis was performed to quantify the decay of response amplitudes across each burst. As such, for each condition and for each burst, we fit the sequence of peak-to-peak response amplitudes to both a single-exponential (equation 2) and a double-exponential (equation 3) decay function:

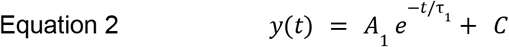

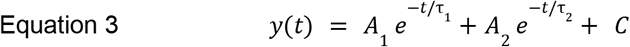

where *A*_1_ and *A*_2_ denote the magnitudes of the fast and slow decaying components, τ_1_ and τ_2_ their corresponding decay time constants, and *C* the asymptotic steady-state offset. Model fitting was performed using nonlinear least-squares optimization (MATLAB lsqcurvefit).

To ensure stable and physiologically meaningful fits, parameters were initialized with reasonable starting values and constrained within predefined bounds. Amplitudes were constrained to be non-negative, decay time constants were restricted to lie within a physiologically plausible range (5-500 ms for the single exponential and 5-2000 ms for the slower component of the double exponential). 2000 ms was considered as the cutoff as bursts lasted no longer than 2 seconds. The constant offset term was allowed to vary freely. For each burst, both single- and double-exponential fits were compared using the Akaike Information Criterion (AIC) and Bayesian Information Criterion (BIC). When the double-exponential model provided a better fit, τ_1_ and τ_2_ from that model were retained; otherwise, the single τ_1_ from the single-exponential model was used. As there were occasional instances where a mix of single- and double-exponential fits were used across conditions, the effective decay time constant (τ_*eff*_) was also calculated to ensure fair comparison (given only one term τ_*eff*_ = τ_1_).

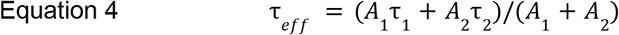

To visualize decay shapes across participants, fitted curves were corrected for the estimated offset term and normalized to the first pulse such that the only variable component between curves was the decay time constant. For double-exponential fits, the amplitude ratio *A*_1_ /*A*_2_ was additionally computed for each burst to assess the relative contribution of the fast and slow components. These ratios were then bootstrapped by resampling with replacement and averaging across iterations. The resulting distribution was used to determine if the overall decay curve was driven primarily by the initial fast decay component (i.e., whether the ratio’s confidence interval remained above zero).

### In-silico model of post-activation depression

To test the impact of volition on modulating post-activation depression, we built a model of afferent fibers, supraspinal fibers, and motoneurons in Python 3.12 using NEURON 8.2^33^.

#### Motoneuron model

We modeled a pool of 100 integrate and fire motoneurons. The model includes an excitatory input current (*E*(*t*)) with an exponential decay constant (τ_*E*_), described in Equation 5, and a leaky integrator membrane state variable (*M*(*t*)) described in Equation 6.

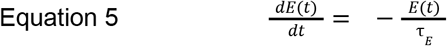

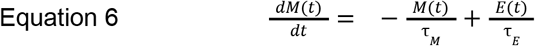

When the value of *M*(*t*) surpasses 1, the cell fires an action potential and the value of *M*(*t*) returns to 0. We used a modified version of the integrate and fire neuron in which, regardless of the value of *M*(*t*), the cell cannot fire again until a set refractory period has elapsed after the time of the firing.

We fit the parameters of the integrate and fire model to reflect the dynamics of motoneuron responses. Specifically, we set the value of τ_*E*_ and τ_*M*_ (τ_*E*_=0.25ms, τ_*M*_=6ms) to reflect an EPSP of approximately 15 ms, as has been shown experimentally^89^. We set the refractory period of each neuron to be drawn from a normal distribution with a mean of 15 ms, resulting in a mean maximum firing rate of 66Hz which is biophysically-reasonable for a sustained hold^90^.

#### Inputs into motoneurons

Each motoneuron received input from all Ia afferents (N=30) and all supraspinal fibers (N=20). SCS frequency determined the rate at which Ia afferent fibers fired (80Hz), thereby modeling the experimental result that afferent fibers are the main target for recruitment SCS pulses^33^. The number of Ia afferents, and therefore the number of SCS inputs, was set to produce small, consistent evoked responses in the rest condition, akin to those produced experimentally by slightly-suprathreshold stimulation. Each pulse of stimulation was modeled to activate the afferents synchronously, an assumption which has been experimentally verified, although the success of each pulse of stimulation was variable due to post-activation depression, as described in more detail in the section below.

The number of supraspinal fibers was determined based on a percentage of the number of supraspinal fibers in the intact model (N=300), thereby simulating the effect of a lesion due to stroke^33^. Each supraspinal fiber was modeled as a Poisson process with a mean firing rate, which was modified to match the level of volitional input. To determine the correct firing rate per volitional condition (10% and 25% of the MVC), we first estimated the maximum firing rate of the motoneuron population. We then set the supraspinal drive frequency to correspond to the production of 10% and 25% of this maximum firing rate (we determined these parameters to be 24Hz and 60Hz respectively). The firing rate of the supraspinal fibers was set to 0Hz during the rest condition.

The synaptic weight of both supraspinal and SCS inputs followed a gamma distribution with the same mean weight. The parameters of this distribution were chosen so that the motoneuron population exhibited flexible recruitment, with motoneurons requiring a variable number of inputs to reach firing threshold, and saturated recruitment with full network activation.

#### Model of post-activation depression

In order to model post-activation depression, we adapted a previously-validated model of spike failure into our model of spinal circuitry^66^. In this model, the two mechanisms by which an SCS-evoked spike in the Ia afferents can fail to reach the motoneuron are failure at the axonal and synaptic level.

First, to model the axon, we set a probability *p* that the spike succeeds in the axon. For each pulse of SCS at *t*_*spike*_ in the axon of fiber *j*, we drew a random uniform value *s* between 0 and 1. The spike would only succeed in the axon if *s* < *p*_*a*_.

To model the synapse, we tracked *n*_*j*_ (*t*), the number of available vesicles at time *t* in the synapse of fiber *j*, with *n*_*j*_ (0) = 3 for each synapse at the start of the simulation *t* = 0. We also set *p*_*r*_, the probability that a vesicle is released given a spike. If the spike at *t*_*spike*_ succeeded in the axon of fiber *j*, the probability of the spike succeeding in the synapse was determined by Equation 7:

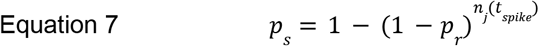

This equation states that the probability of a spike succeeding in the synapse is the complement of the probability that no vesicles from that synapse are released. Again, we next chose a random variable *r* from a uniform distribution between 0 and 1, and record a successful synaptic spike if *r* < *p*_*s*_. The success at the synapse, therefore, relies on the number of vesicles available; if there are no vesicles remaining, *p*_*s*_ = 0 and the spike cannot succeed in the synapse.

If a spike does succeed in both the synapse and the axon, the number of available vesicles *n*_*j*_ (*t*) is decremented by one. To model the reuptake of the vesicles to the synapse after a spike, we draw a random exponential value with mean τ_*r*_ after each successful spike. Once this wait time has elapsed, the number of available vesicles *n*_*j*_ (*t*) is incremented by one. A successful spike in the axon and synapse is received by the motoneuron as an excitatory Ia afferent input, as described above.

To utilize this model to explore the role of corticospinal drive in modulating post-activation depression, we modified two key parameters. To test Hypothesis 2, in which we posited that corticospinal drive may be increasing the probability of axonal success, we increased *p*_*a*_ relative to the level of volitional drive (rest: *p*_*a*_ = 0. 3, 10% effort: *p*_*a*_ = 0. 5, 25% effort: *p*_*a*_ = 0. 8). To test Hypothesis 3, in which we tested if corticospinal drive could be increasing the rate of reuptake, we decreased τ_*r*_ relative to the level of volitional drive, thereby producing quicker reuptake timescales (rest: τ_*r*_ = 100, 10% effort: τ_*r*_ = 68, 25% effort: τ_*r*_ = 20). Besides these changes, we maintained the same parameters across the simulated conditions in order to evaluate the impact of specific hypothesized parameters.

#### Estimation of EMG signal

To estimate the EMG signal from the motoneuron firing rates, we developed an approach based on previously-described methods^36,68,91^. First, we start with the boolean firing matrix *F*, in which each row is a motoneuron number and each column is the time sample, such that *F*_*i,t*_ = 1 if the motoneuron *i* fires at time *t*. For a given motoneuron *i*, we create a motor unit action potential (MUAP) by drawing a length *L*_*i*_ and amplitude *A*_*i*_ from a normal distribution (*L*_*i*_ ~ *N*(7*ms*, 1*ms*)^92^, *A*_*i*_ ~ *N*(1*au*, 0. 2*au*)). Using these parameters, we defined an amplitude envelope *a*_*i*_ (*n*), where n is the time step in the MUAP and the amplitude remains constant at *A*_*i*_ for the first 40% of the duration, and then decreases to ⅓ of the initial amplitude for the last 60% of the duration of the MUAP. Using a base *b* = 1. 05, we generated the sine-wave vector *s*_*i*_ (*n*) using Equation 8:

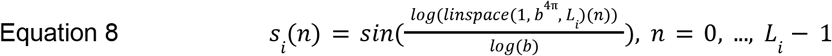

Finally, we used these values to compute the MUAP vector *M*_*i*_ (*n*) for motoneuron *i*, where *n* is the time steps in *L*_*i*_, using Equation 9:

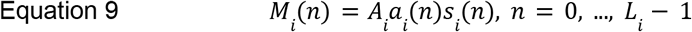

At a given spike at time *t*, we constructed the final EMG signal, *E*(*t*), by summating the underlying MUAPs. *E*(*t*) is initialized to be a zero vector of length *T*, the duration of the simulation. If *F*_*i,t*_ = 1, we utilized Equation 10 to modify *E*(*t*) to reflect the MUAP.

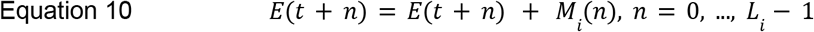

By performing this operation for all time points *t* and all motoneurons *i*, we were able to generate an estimate of the EMG signal that reflects the summation of biophysically-reasonable underlying MUAPs^92^.

With this estimate of the EMG signal, we followed the protocol described above to calculate the peak-to-peak amplitude of evoked responses and fit the depression to an exponential curve. Due to the relative simplicity of the model, we did not consider a double exponential function in fitting data from the model, but rather fit the decay in amplitude to a single exponential function across all experimental conditions.

### Passive vs active movement

Participants were seated in the robotic isokinetic machine described previously (HUMAC NORM, CSMi) with the hand secured to the manipulandum and the elbow aligned with the device’s axis of rotation. The machine guided the arm through approximately 30° of cyclic elbow flexion and 30° of elbow extension, centered around an elbow angle of 90°. Joint limits were occasionally adjusted to accommodate participant comfort or available range of motion. Surface EMG electrodes were placed over the biceps and triceps.

Two isokinetic movement conditions were tested. In the passive condition, the HUMAC moved the arm through the flexion/extension cycle at a fixed speed of 10°/s while participants remained relaxed. In the active condition, participants attempted to volitionally flex or extend their arm in the same direction as the HUMAC’s movement while it remained at the same fixed speed. Flexion and extension tasks were separated to minimize fatigue and cognitive load; for example, during the flexion task participants actively flexed during the flexion phase (30° to 120°) and relaxed during the extension phase (120° to 30°). During each condition, low-frequency suprathreshold SCS (5-10 Hz) was delivered continuously through a single contact, selected to evoke clear biceps or triceps reflex responses. Each task consisted of six repetitions of the flexion/extension cycle per condition.

Stimulation-triggered responses were extracted for every SCS pulse across repetitions. Responses that exceeded baseline threshold and fell within the tested phase of movement were retained and tagged with the corresponding elbow angle at the time of stimulation. For each task, condition, and muscle, responses were binned by their tagged elbow angle, and the mean peak-to-peak response amplitude within each bin was calculated. To account for overall increases in excitability during active movement, reflex amplitudes within each bin were normalized to the mean amplitude of the bin with the largest responses. A linear fit was then applied to the mean response of each angle bin to quantify the amplitude-angle relationship (slope), which served as an index of how reflex amplitude changed with muscle length under passive versus active movement.

### EMG analysis

Surface electromyography (EMG) was recorded using either wireless sensors (Avanti Trigno, Delsys Inc.) or high-density EMG (HDEMG) grids (TMSi, The Netherlands), depending on the requirements of each task; in some experiments, both systems were used simultaneously. Delsys sensors sampled EMG at 1926 Hz with onboard 20–800 Hz hardware bandpass filtering, and signals were resampled at 8000 Hz through a National Instruments DAQ for synchronization. Flexible HDEMG grids of either 8×8 or 8×4 channels were placed over the relevant muscle groups (e.g., biceps and triceps; forearm flexors and extensors) and recorded in a monopolar configuration using a TMSi Saga 64+ amplifier at 2 kHz. Conductive gel was applied to reduce skin-electrode impedance and a reference electrode was placed over the lateral epicondyle. All EMG data was time-aligned and appropriately resampled to ensure synchronization across devices.

#### Root-mean-square (RMS) amplitude

EMG signals from each recorded muscle were band-pass filtered between 10 and 200 Hz using a zero-lag 4th-order digital Butterworth filter and then notch-filtered at stimulation harmonics using a zero-lag 4th-order infinite impulse response (IIR) band-stop filter. For MVC trials, RMS was calculated across each 5 second contraction and then averaged for further analysis. For force-control tasks, RMS was calculated using a moving-average window (500ms). RMS vectors across repetitions were then averaged and sampled at specific timepoints to capture mean muscle activation during the rising and falling phases of force production. For planar reaching, RMS was calculated across the reaching phase and then averaged for further analysis.

When relevant, polar plots were generated using the mean RMS amplitudes for both SCS OFF and SCS ON. Each recorded muscle was assigned to a radial axis while the radial magnitude reflected mean RMS activation. The change in the proportion of agonist RMS relative to total multi-muscle RMS between SCS OFF and SCS ON conditions was additionally calculated and annotated on the plot.

#### Co-contraction index

EMG signals from the biceps and triceps or brachioradialis and triceps were band-pass filtered between 10 and 200 Hz using a zero-lag 4th-order digital Butterworth filter and then notch-filtered at stimulation harmonics using a zero-lag 4th-order IIR band-stop filter. Co-contraction between these flexor and extensor muscles was then quantified using agonist-antagonist activation ratios. For each participant and stimulation condition (SCS ON and SCS OFF), the RMS (MVC) or the moving average RMS (force-control) was used in the following equation to quantify changes in muscle co-contraction due to SCS as the percentage change in agonist-antagonist activation ratios (i.e., AG./ANTAG.), computed as the ratio of values during SCS ON relative to SCS OFF (i.e., SCS ON/SCS OFF):

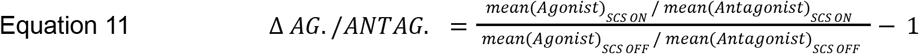

#### Intermuscular coherence

EMG signals from the biceps and triceps were band-pass filtered between 10 and 200 Hz using a zero-lag 4th-order digital Butterworth filter and then notch-filtered at stimulation harmonics using a zero-lag 4th-order IIR band-stop filter. Intermuscular coherence was then computed between pairs of muscles (e.g. biceps and triceps or biceps and brachioradialis) during the elbow flexion fine force-control task. Preprocessed EMG signals were analyzed using Welch’s method (mscohere in MATLAB) with 0.5s Hanning windows and 50% overlap. For each repetition and condition (SCS OFF and SCS ON), coherence spectra were first estimated separately and then pooled by summing the corresponding auto- and cross-spectra across repetitions (pwelch and cpsd respectively) before recalculating coherence as shown in Equation 12.

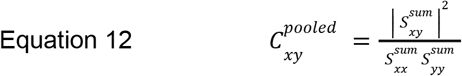

The 95% confidence limit for coherence was derived from the effective number of Welch segments (*N*) summed across repetitions (*K*_*pooled*_) (Equation 13)

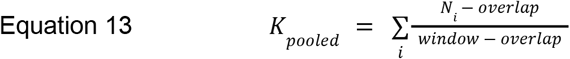

and calculated as:

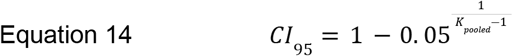

Beta-band coherence was quantified as the area of coherence above this 95% confidence limit and between 13-30hz.

#### Stimulation-triggered responses

EMG signals from each recorded muscle were band-pass filtered between 10 and 500 Hz using a zero-lag 2nd-order digital Butterworth filter. EMGs were also manually inspected after preprocessing and filtering. Muscles that remained excessively noisy were excluded from further analysis. Stimulation pulses recorded on the National Instruments DAQ were used to identify individual stimulation time-points. In the event that triggers were not properly recorded, stimulation time-points could also be determined from artifacts in the EMG, particularly in the deltoids. For each stimulation event, a stimulus-triggered EMG window was extracted. Window durations were manually adjusted to ensure that the full evoked response was captured. In some cases, the start of the window was shifted (typically 5-10 ms) to avoid including the stimulation artifact. For the passive vs. active experimental protocol and for all protocols involving TMS, a window preceding each stimulation was also extracted to estimate baseline muscle activity. This was then used to determine whether the subsequent window contained a true stimulation-evoked response. If the maximum absolute value of a stimulation-triggered window exceeded the average rectified background activity by 2 standard deviations (passive vs active) or by a more rigorous 4 standard deviations (TMS+SCS paired-pulse protocol), a response was considered real. In the event of TMS-related protocols, windows that failed to contain a real-response were zeroed. The peak-to-peak amplitude of each stim-triggered response was then used for further analysis.

#### Motor unit decomposition

High density surface electromyography (HDEMG) was recorded from forearm muscles in SCS05, SCS07 and SCS08. Participants were asked to perform a steady grip force modulation using a handheld dynamometer wherein they had to sustain 20% of their maximum voluntary contraction for 1 minute. HDEMG recordings were decomposed into individual motoneurons spike trains using DEMUSE tool software which exploits the convolution kernel compensation (CKC) method^93^. The results of this automatic decomposition were manually edited following the standard procedure already described^94^. Only motoneurons with pulse-to-noise ratio ≥ 25 dB were selected for the analysis.

### Quantification and statistical analysis

All statistical analyses were performed offline in MATLAB (R2025a). Exact sample sizes (n) and what n represents (e.g., repetitions, bursts, or participants) are reported in the figure legends. Data visualizations include boxplots (median ± IQR), bar and line plots summarizing individual participant metrics, and bootstrap-derived confidence intervals for group-level effects for which individual repetitions or participant-level values are overlaid when appropriate.

For all comparisons of SCS OFF vs. SCS ON, and for passive vs. active conditions, participant-level metrics (e.g., MVC torque, RMSE during fine force control, normalized reflex slopes, EMG activity) were first computed within each participant. Group-level inference was then performed using nonparametric bootstrap resampling: participant-level mean differences were resampled with replacement (10,000 iterations) to generate a distribution of the group mean, from which the 95% confidence interval (CI) was taken. CIs excluding zero were interpreted as statistically significant when testing the null hypothesis of no change between conditions. When directly comparing SCS OFF vs SCS ON as is done for MVC torque in Figure 7, this same resampling framework was used and asterisks denote cases wherein CIs indicate significant differences in the distribution between conditions.

For other within-participant trial-level statistical comparisons (e.g., TMS+SCS reflex conditioning by ISI and decay time-constants by effort level), nonparametric tests were used. Conditioned reflex amplitudes were compared using Kruskal-Wallis tests with Dunnett-type post hoc comparisons against the no-conditioning baseline, and effect sizes were reported as eta-squared (η^2^). For post-activation depression, fitted decay time-constants were compared using Kruskal-Wallis tests with Tukey-Kramer corrections. Statistical thresholds were defined as \**p* < 0.05, \*\**p* < 0.01, and \*\*\**p* < 0.001 unless otherwise noted.

Intermuscular coherence was quantified as described in the EMG processing section. Statistical significance of coherence values was determined using the 95% confidence threshold derived from the pooled number of independent spectral segments, and pooled coherence exceeding this threshold was considered significant. For later polar-plot visualizations, EMG RMS activity reflects the mean RMS across repetitions, and changes in muscle recruitment are summarized as the percent change in primary agonist to total EMG RMS.

All analyses utilized data which met manual quality-control criteria for EMG signal quality, stimulation delivery, and task compliance (e.g., trials with incorrect stimulation parameters, compensatory movements, or inadequate relaxation during passive/rest conditions were excluded/repeated). Trial condition orders were randomized where feasible (e.g., interstimulus intervals, passive vs. active blocks, etc.). All statistical tests, effect sizes, confidence intervals, and n definitions are reported in the figures or figure legends.

## ADDITIONAL RESOURCES

Participants were recruited for clinical trial NCT04512690, further information can be found at: https://clinicaltrials.gov/study/NCT04512690.

## SUPPLEMENTAL FIGURES

**Figure S1.**
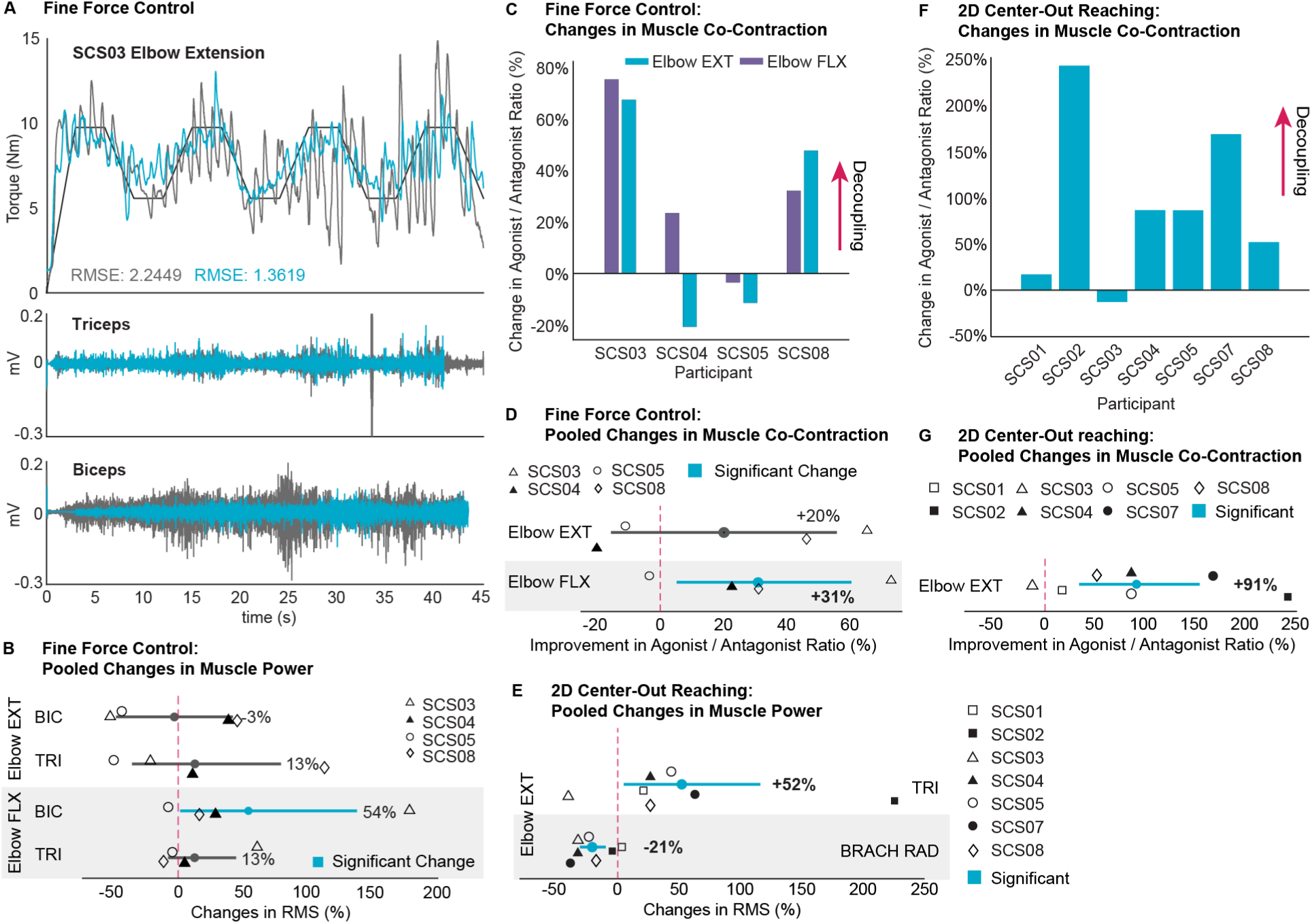
Spinal cord stimulation improves muscular coordination, related to Figure 1. **(A)** Example fine force control during 20–35% MVC elbow extension (SCS03). Torque traces across time are shown for SCS OFF (grey) and SCS ON (blue), n=1 per condition. RMSE from the target reported for each. Corresponding triceps and biceps EMG show reduced overall activity and a temporal shift in bicep activation toward the off periods of triceps during SCS ON. **(B)** Group-level changes in muscle power (EMG RMS) for the triceps (TRI) and biceps (BIC) during 20–35% MVC fine-force control. Percent differences were calculated per participant between mean SCS ON and SCS OFF (extension: n=1–3, flexion: n=1–3 per condition) and bootstrapped to obtain 95% CIs. **(C)** Per-participant changes in co-contraction during 25–30% MVC fine-force control, quantified as the mean agonist/antagonist EMG ratio (see methods). Increases indicate greater decoupling with SCS ON. **(D)** Group-level changes in co-contraction, quantified as the agonist/antagonist EMG ratio (see methods), during 20–35% MVC fine-force control. Percent differences were calculated per participant between mean SCS ON and SCS OFF (extension: n=1–3, flexion: n=1–3 per condition) and bootstrapped to obtain 95% CIs. Increases indicate improved decoupling with SCS ON. **(E)** Group-level changes in muscle power (EMG RMS) for the triceps (TRI) and brachioradialis (BRACH RAD) during 2D reaching. See (B). **(F)** Per-participant changes in co-contraction during 2D reaching. See (C). **(G)** Group-level changes in co-contraction during 2D reaching. See (D). *Panels B, D, E, and G: Participant-level mean percent differences were resampled with replacement (10,000 iterations) to estimate the group-level mean and 95% CI. CIs excluding zero (blue) indicate significant changes; the mean percent change is shown for each interval*.

**Figure S2.**
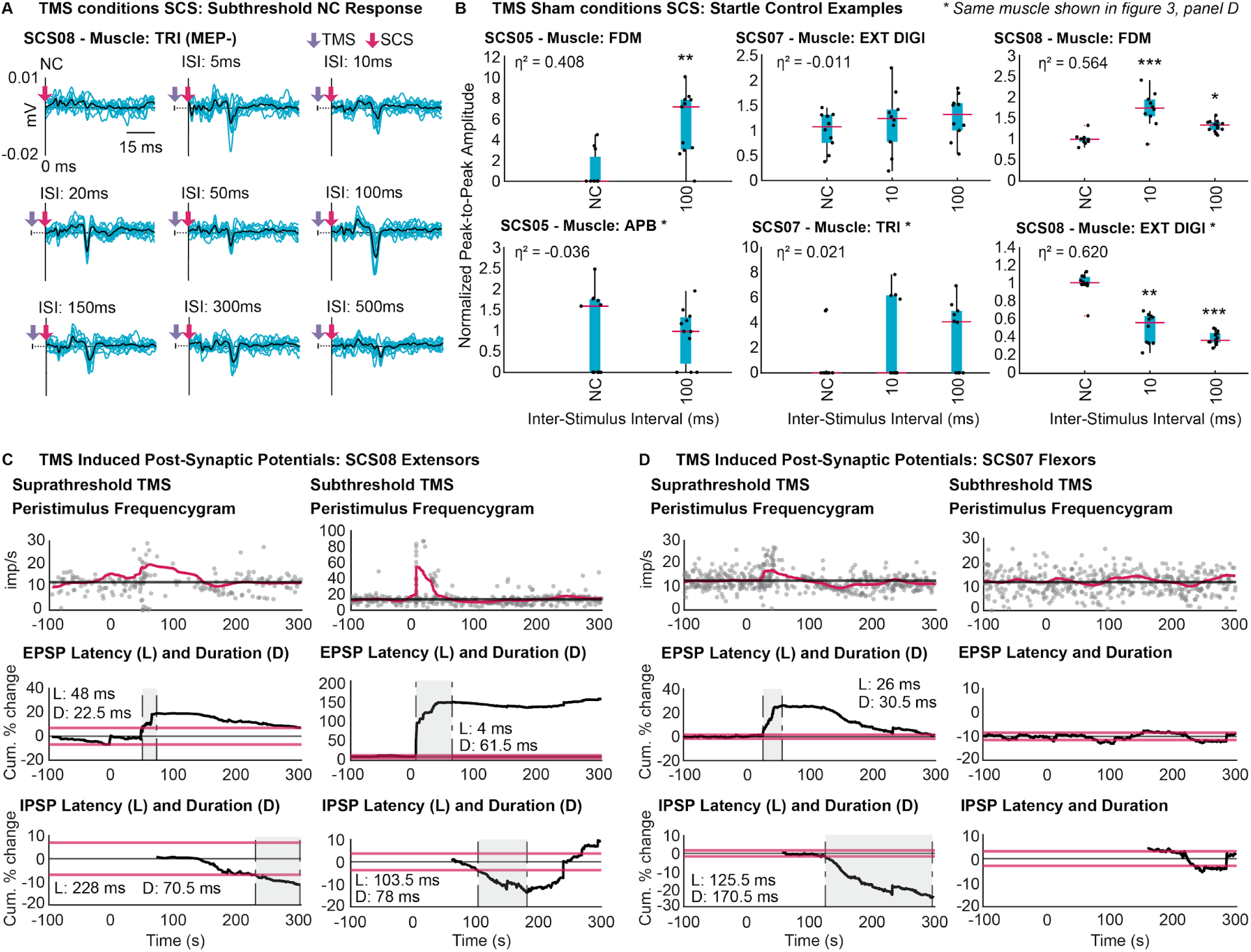
Residual CST, not RST, responsible for presynaptically facilitating reflexes at long ISIs, related to Figure 3. **(A)** Extra example showing that even when SCS does not evoke a response in the no-conditioning (NC) condition, subthreshold TMS can cause the emergence of a response at both short and long ISIs, even in an MEP-muscle (SCS08: TRI). Individual responses (blue); mean response (black). **(B)** TMS sham control: Startle-evoked responses during sham TMS in representative muscles from the participants who underwent this protocol (SCS05, SCS07, SCS08). Muscles that showed significant facilitation under sham conditions were limited to SCS05 ECR and FDM, and SCS08 FCR. Starred panels indicate the same muscles shown in Figure 3, where TMS facilitated SCS-evoked responses. Boxplots (median ± IQR, whiskers = 1.5×IQR) show normalized peak-to-peak SCS-evoked response amplitudes across NC, 10ms ISI, and 100ms ISI with overlaid dots (n=10–13 per ISI). *Kruskal–Wallis with Dunnett-type post hoc versus NC; η*^*2*^ *effect size. *p<0*.*05, **p<0*.*01, ***p<0*.*001*. **(C–D)** TMS-evoked postsynaptic potentials: Peristimulus frequencygrams (PSFs) from motor-units (MU) in **(C)** SCS08 extensors and **(D)** SCS07 flexors during suprathreshold (left column) and subthreshold (right column) TMS (SCS08 supra: n=2 MU; SCS08 sub: n=1 MU; SCS07 supra: n=3 MU; SCS07 sub: n=4 MU). Note, SCS07 flexors showed no detectable PSPs following subthreshold TMS. Top panel: firing probability aligned to TMS onset (grey dots: individual instantaneous firing rates; pink: moving average; black: mean background firing rate). Bottom panels: cumulative-sum analyses used to quantify excitatory (EPSP) and inhibitory (IPSP) postsynaptic potentials, with labeled latency (L) and duration (D). Pink bounding boxes: baseline thresholds; black: cumulative sum trace; shaded region: EPSP or IPSP period.

**Figure S3.**
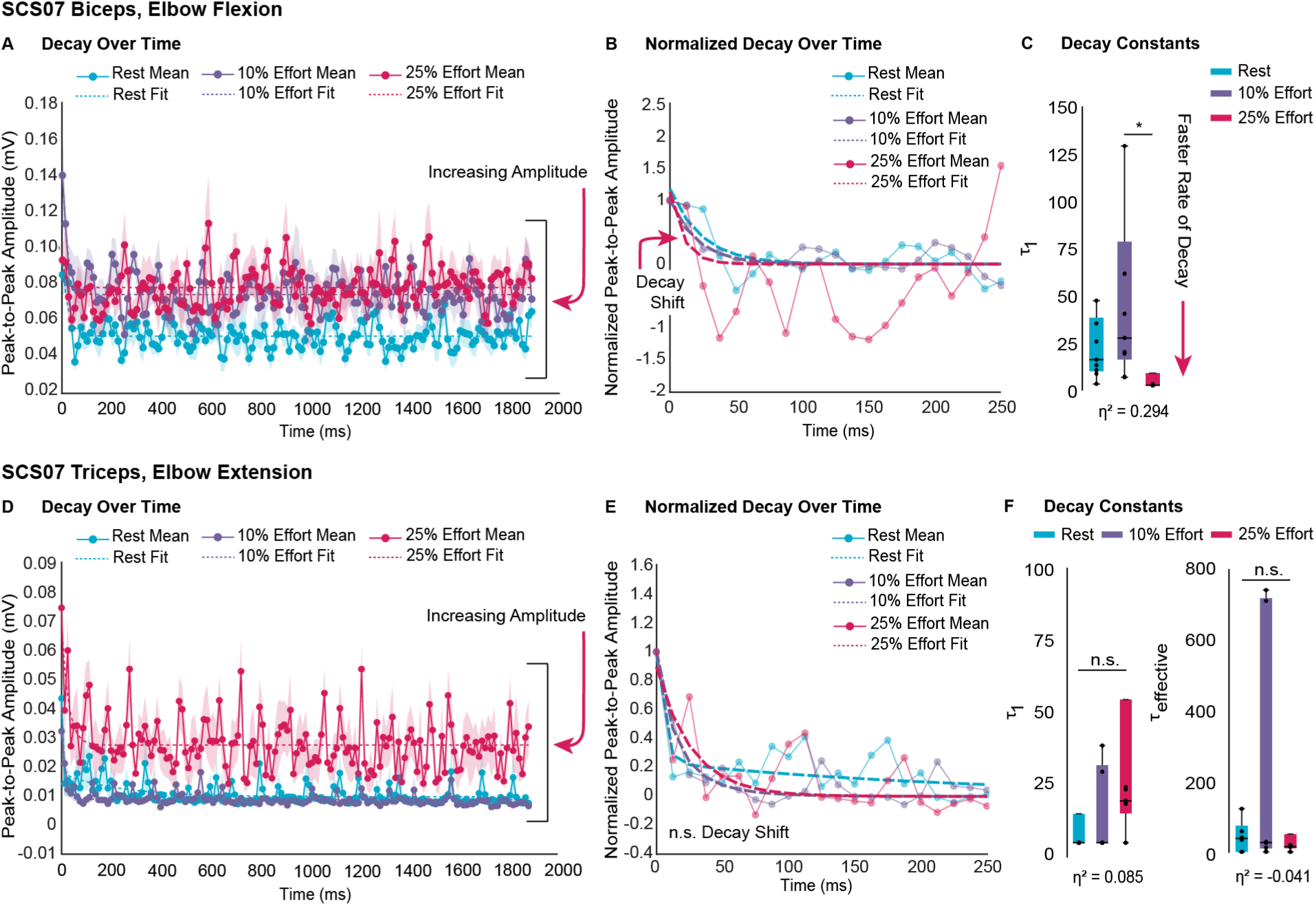
In vivo: volitional modulation of post-activation, examples from SCS07, related to Figure 5. **(A–C)** SCS07, elbow flexion, biceps. **(A)** Peak-to-peak amplitude across 80hz burst (mean (solid line) ± SEM; single exponential fit (dashed line); n=9 bursts). **(B)** Normalized peak-to-peak amplitude across 80hz burst (mean (solid line); single exponential fit (dashed line); n=9 bursts). **(C)** Boxplots (median, IQR, whiskers = 1.5×IQR) summarize fitted decay time constants (τ_1_) with overlaid dots (n=9 bursts). **(D–F)** SCS07, elbow extension, triceps. **(D)** Peak-to-peak amplitude across 80hz burst (mean (solid line) ± SEM; exponential fit (dashed line); double-exponential used if two-term model favored; n=9 bursts). **(E)** Normalized peak-to-peak amplitude across 80hz burst (mean (solid line); exponential fit (dashed line); double-exponential used if two-term model favored; n=9 bursts). **(F)** Boxplots (median, IQR, whiskers = 1.5×IQR) summarize fitted decay time constants (τ_1_, τ_*eff*_) with overlaid dots (n=9 bursts). *Rest: blue; 10% effort: purple; 25% effort: pink. Panels C, F: Kruskal-Wallis with Tukey-Kramer post hoc, *p<0*.*05, **p<0*.*01, ***p<0*.*001. Effect size reported as η*^*2*^.

## SUPPLEMENTAL TABLES

**Table S1.**
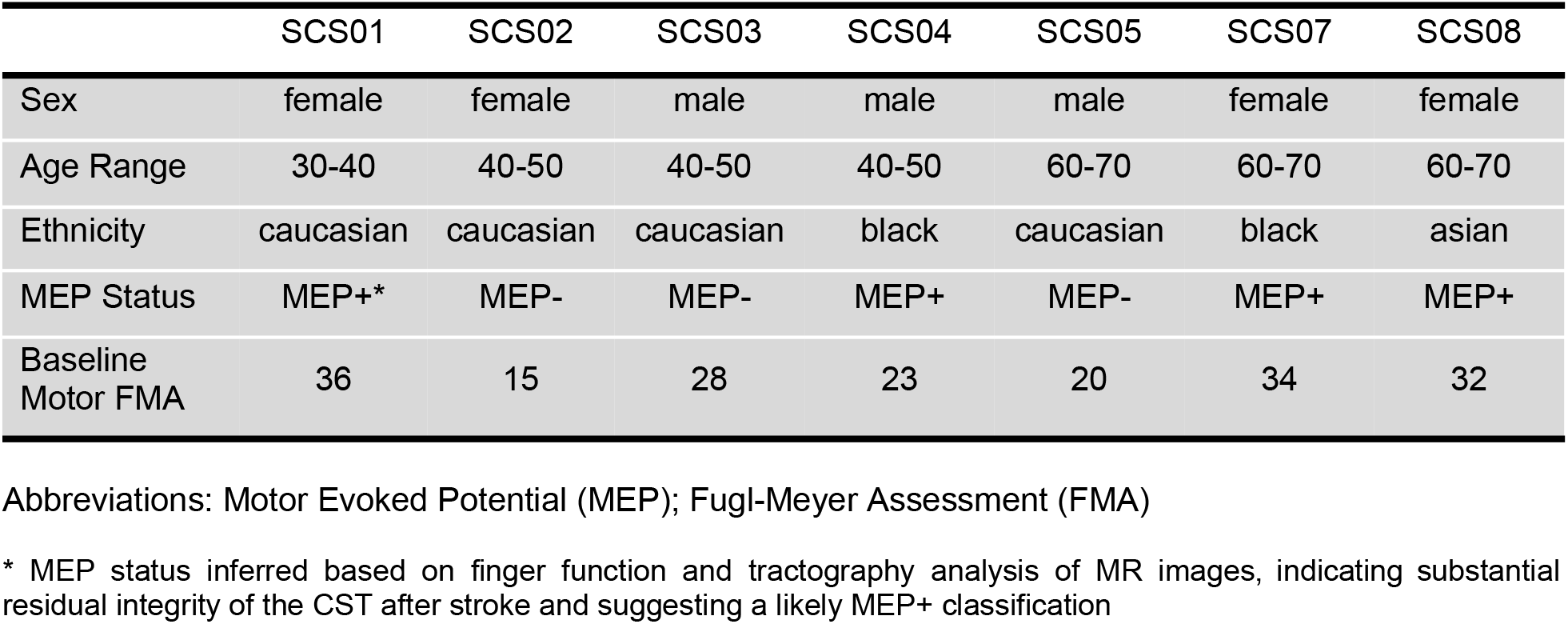
Participant Characterization.

|  | SCS01 | SCS02 | SCS03 | SCS04 | SCS05 | SCS07 | SCS08 |
| --- | --- | --- | --- | --- | --- | --- | --- |
| Sex | female | female | male | male | male | female | female |
| Age Range | 30-40 | 40-50 | 40-50 | 40-50 | 60-70 | 60-70 | 60-70 |
| Ethnicity | caucasian | caucasian | caucasian | black | caucasian | black | asian |
| MEP Status | MEP+* | MEP- | MEP- | MEP+ | MEP- | MEP+ | MEP+ |
| Baseline Motor FMA | 36 | 15 | 28 | 23 | 20 | 34 | 32 |

Abbreviations: Motor Evoked Potential (MEP); Fugl-Meyer Assessment (FMA)
\* MEP status inferred based on finger function and tractography analysis of MR images, indicating substantial residual integrity of the CST after stroke and suggesting a likely MEP+ classification

**Table S2.** Experimental Participation.

|  | SCS01 | SCS02 | SCS03 | SCS04 | SCS05 | SCS07 | SCS08 |
| --- | --- | --- | --- | --- | --- | --- | --- |
| Maximal Volitional Contractions |  |  |  |  |  |  |  |
| Elbow Ext. | yes | -- | yes | yes | yes | yes | yes |
| Elbow Flx. | yes | yes | yes | yes | yes | yes | yes |
| Grip | yes | yes | yes | yes | yes | yes | yes |
| Fine Force Control |  |  |  |  |  |  |  |
| 20-35% Elbow Ext. | -- | -- | yes* | yes* | yes* | yes | yes* |
| 20-35% Elbow Flx. | -- | -- | yes* | yes* | yes* | yes | yes* |
| 20-35% Grip | -- | -- | -- | yes | yes | -- | yes |
| 35-50% Elbow Ext. | -- | -- | yes | yes | yes | yes | yes |
| 35-50% Elbow Flx. | -- | -- | yes | yes | yes | yes | yes |
| 35-50% Grip | -- | -- | -- | yes | yes | -- | yes |
| 2D Center-Out |  |  |  |  |  |  |  |
| Elbow Ext. | yes* | yes* | yes* | yes* | yes* | yes* | yes* |
| TMS |  |  |  |  |  |  |  |
| SCS c. TMS | -- | -- | -- | yes | yes | yes | -- |
| TMS c. SCS | -- | yes | yes | yes | yes | yes | yes |
| TMS Startle Control | -- | -- | -- | -- | yes | yes | yes |
| TMS-PSF | -- | -- | -- | -- | yes | yes | yes |
| Post-Activation Depression |  |  |  |  |  |  |  |
| Elbow Ext. | -- | -- | -- | -- | -- | yes | yes |
| Elbow Flx. | -- | -- | -- | -- | -- | yes | yes |
| Passive vs. Active Reflexes |  |  |  |  |  |  |  |
| Elbow Ext. | -- | -- | yes | yes | -- | yes | yes |
| Elbow Flx. | -- | -- | yes | yes | -- | yes | yes |

Abbreviations: extension (ext.); flexion (flx.); Spinal Cord Stimulation (SCS); Transcranial Magnetic Stimulation (TMS); conditions (c.); Peristimulus Frequencygram (PSF)
\* EMG recorded and analyzed in manuscript

## Notes

### Competing Interest Statement

Authors D.J.W., P.G., and M.C. are founders and shareholders of Reach Neuro, a company developing SCS technologies for stroke. E.P. has interest in Reach Neuro due to personal relationships with M.C.
Authors E.S., N.V., D.J.W., L.E.F., P.G., E.P., and M.C. are inventors on patents related to this work. All other authors declare no competing interests.

### Clinical Trial

NCT04512690

### Author Declarations

IRB STUDY19090210 of University of Pittsburgh gave ethical approval for this work.

### Summary of Updates

The legend for Figure 7 was revised.

